# Sex-dependent placental mQTL provide insight into the prenatal origins of childhood-onset traits and conditions

**DOI:** 10.1101/2022.10.04.22280695

**Authors:** William Casazza, Amy M. Inkster, Giulia F. Del Gobbo, Victor Yuan, Fabien Delahaye, Carmen Marsit, Yongjin P. Park, Wendy P. Robinson, Sara Mostafavi, Jessica K Dennis

## Abstract

Molecular quantitative trait loci (QTL) allow us to understand the biology captured in genome-wide association studies (GWAS). The placenta regulates fetal development, and shows sex differences in DNA methylation. We therefore hypothesized that placental methylation QTL (mQTL) explains variation in genetic risk for childhood-onset traits, and does so differentially by sex. We analyzed 411 term placentas from two studies and found 49,252 methylation (CpG) sites with methylation QTL (mQTL) and 2,489 CpG sites with sex-dependent mQTL. All mQTL were enriched in regions active in prenatal tissues that typically affect gene expression. All mQTL were enriched in GWAS results for growth- and immune-related traits, but male- and female-specific mQTL were more enriched than cross-sex mQTL. mQTL colocalized with trait loci at 777 CpG sites, with 216 (28%) specific to males or females. Overall, mQTL specific to male and female placenta capture otherwise overlooked variation in childhood traits.

## Introduction

GWAS findings hold valuable clues about trait mechanisms.^1,2^ Deciphering these clues, however, requires additional data on gene regulation, as over 90% of SNPs identified in GWAS lie in gene regulatory regions, as opposed to in the protein-coding gene region itself.^3,4^ Molecular quantitative trait locus (molQTL) analysis is a powerful strategy to interpret the gene regulatory functions of GWAS SNPs. Under the umbrella of molQTL, expression quantitative trait loci (eQTL) are the most widely studied, and they are enriched for GWAS loci relative to other SNPs matched by minor allele frequency (MAF).^5^ However, only 43% of eQTL share the same causal variant (colocalize) with a GWAS locus,^5^ and up to 77% of eQTL in linkage disequilibrium (LD) with a trait-associated SNP are shared across more than one tissue.^6^ As a result, the majority of GWAS loci have either no known effects on expression, or their relationship with traits is clouded by their broad effects on gene expression across tissues.^5,7^ Moreover, few new eQTL are being discovered in the most widely used eQTL resource, GTEx, even as sample sizes exceed 600,^7^ suggesting that eQTL discovery using post-mortem adult tissues is limited.

Therefore, to advance the functional interpretation of GWAS SNPs, we must extend molQTL discovery across molecular traits, tissues, and biological contexts. In this study, we identify DNA methylation QTL (mQTL) in placenta, and additionally focus on mQTL that have different effects in males vs. females. DNAm is an attractive molecular trait for functional interpretation of GWAS results because it can provide insights on the precise molecular mechanism by which GWAS SNPs associate with traits and conditions: through biochemical modification to DNA sequence at a CpG site.^8^ In addition, variation in DNAm is genetically influenced. In blood, the tissue in which DNAm is most widely studied, 21% of the variation of DNAm is explained by additive genetic variation in cis (i.e., via mQTL).^9^ Importantly, mQTL provide information on gene regulation beyond what is provided by eQTL. For example, mQTL cover roughly twice as many genes as eQTL in blood,^5,9,10^ and in one study measuring both mQTL and eQTL in blood in the same participants, only 7% of mQTL with eQTL effects actually mediated the effect of that locus on gene expression, suggesting that the DNAm was regulating a distinct molecular process.^10^

The placenta is of the same genetic make up as the fetus and it is one of the first organs to form during gestation. Throughout pregnancy it is responsible for the exchange of oxygen, nutrients, and hormones between mother and fetus. Despite its central role in human development, however, placenta is under-studied and is not represented in large-scale molQTL resources like GTEx. Several studies have previously investigated placental molQTL and their relationship with complex traits and conditions.^11–16^ In the largest mQTL analysis to date, Delahaye et al.^12^ analyzed 303 placental samples from the the Eunice Kennedy Shriver National Institute of Child Health and Human Development (NICHD) study and characterized a small number (N = 4,342) of strongly associated (i.e., passing a stringent permutation test and quality threshold) mQTL, which were found to overlap two type 2 diabetes loci. Tekola-Ayele et al.^13^ analyzed the same samples with a similar approach and using colocalization analysis, implicated four genes with placental DNAm and gene expression that shared genetic loci with birth weight. While these analyses of the NICHD study demonstrated the relevance of placental molecular traits to postnatal outcomes, the stringent thresholds used to map mQTL means that many mQTL remain unmapped, especially in larger sample sizes. In addition, both Delahaye and Tekola-Ayele analyzed at most two GWAS traits, and the relationship of placental mQTL to multiple postnatal outcomes has yet to be investigated. The Rhode Island Child Health Study (RICHS)^15,17^ is another large study that has collected molecular data from 149 placental samples, but mQTL have not been mapped in the RICHS study, and overall, a more comprehensive study is needed to better understand how placental mQTL affect genome-wide risk of traits.

Additionally, neither the NICHD nor RICHS studies have investigated sex differences in the genetic regulation of placental molecular traits. Oliva et al. recently analyzed sex differences in eQTL across 44 GTEx tissues and found that sex-dependent eQTL (i.e., cross-tissue mQTL with a genotype by sex interaction) were remarkably tissue specific: of 369 sex-dependent eQTL found in at least one tissue, only one was shared by two tissues.^18^ Moreover, they found 74 eQTL in either males or females that colocalized with GWAS loci, 24 of which showed no evidence of colocalization in eQTL computed in all subjects. These results suggest that sex-dependent molQTL yield functional interpretations of GWAS loci beyond what is provided by cross-sex analysis. Analyzing sex-dependent molQTL could be especially important in placenta, since sex is strongly associated with placental molecular traits such as DNAm, even after accounting for cell type proportions,^19,20^ which can bias sex-dependent molQTL analysis.^18^

In this work we identify placental mQTL with a shared effect in males and females (cross-sex mQTL) and mQTL that are modified by sex (sex-dependent mQTL) by meta-analyzing data from 411 term placentas from the NICHD study and RICHS. We then compare cross-sex and sex-dependent mQTL with regards to their genomic location. Finally we quantify the relevance of cross-sex and sex-dependent mQTL to childhood-onset traits and conditions using stratified linkage disequilibrium score regression (S-LDSC)^21–23^ and colocalization analysis.

## Results

### Sex-dependent placental mQTL are distinct from cross-sex placental mQTL

We meta-analyzed mQTL across the NICHD and RICHS studies (Table S1), using MeCS^24^ to account for bias induced by correlation in mQTL effects between studies. Meta-analyzed mQTL effects (or genotype by sex interaction effects in the sex-dependent analysis) were called at a Bonferroni corrected p < 0.05. We defined 6 sets of mQTL (Figure 1, Table 1) which we use in all downstream analyses: (i) cross-sex, which have an effect independent of sex (typically just referred to as “mQTL”); (ii) sex-dependent, which have an effect modified by sex; (iii) male-stratified, which have an effect in male samples; (iv) female-stratified, which have an effect in female samples; (v) male-specific,which have an effect in males that also differs from the effect in females (intersection of sets (ii) and (iii)); and (vi) female-specific, which have an effect in females that differs from the effect in males (intersection of sets (ii) and (iv)).

**Figure 1.**
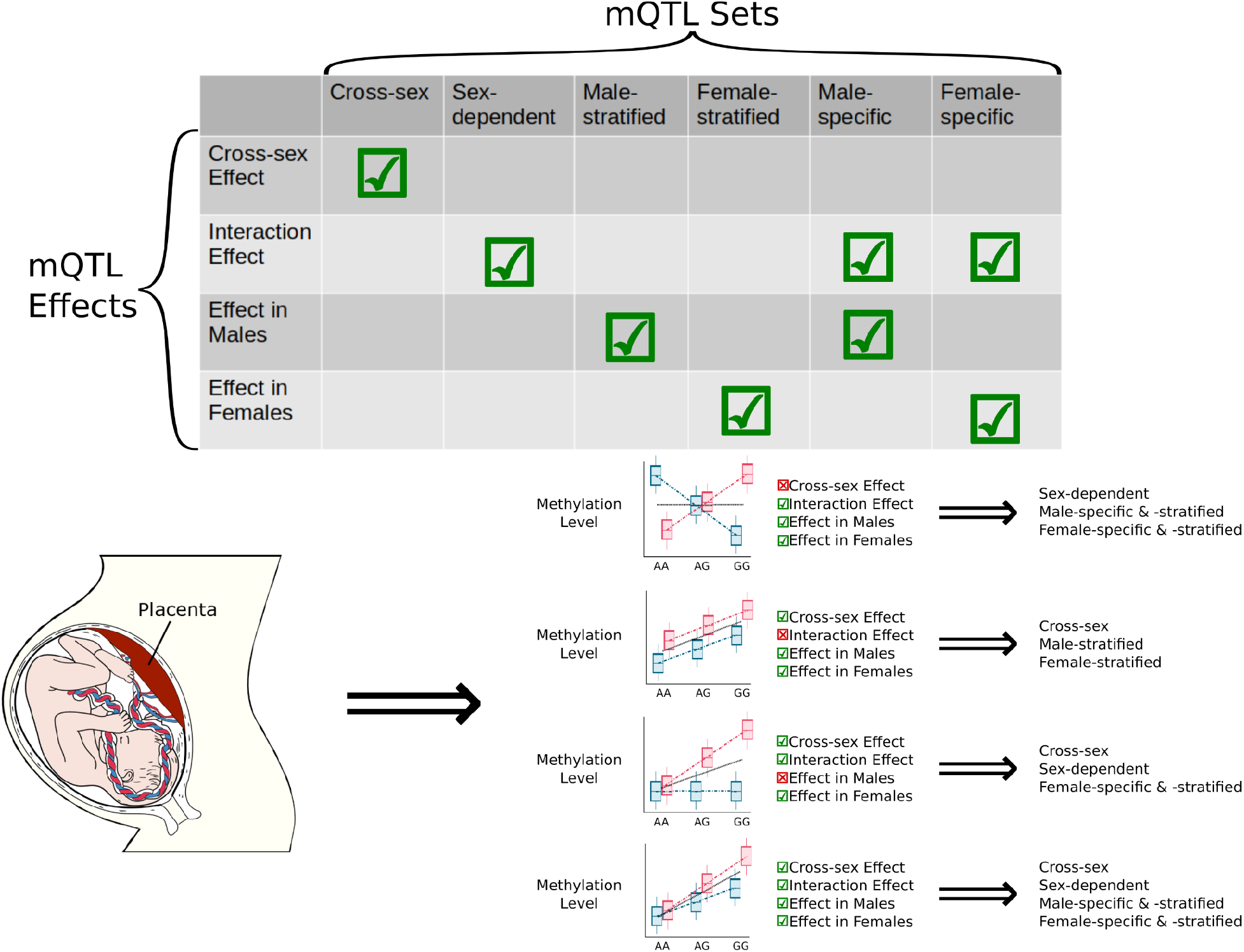
Defining placental mQTL sets from mQTL effects. We defined, (i) cross-sex, which have an effect independent of sex; (ii) sex-dependent, which have an effect that differs between male and female samples (a genotype by sex interaction effect), (iii) male-stratified, which have an effect in males, (iv) female-stratified, which have an effect in females, (v) male-specific, which have an effect in males that also differs from the effect in females, and (vi) female-specific, which have an effect in females that also differs from the effect in males.

**Table 1.** Counts of mQTL, associated CpG sites and corresponding genes in each mQTL set. CpG sites with at least one mQTL were included, and corresponding genes were identified using Illumina’s HumanMethylation450k array annotation.

| mQTL Set | No. mQTL | No. CpG Sites | No. Corresponding Genes |
| --- | --- | --- | --- |
| Cross-sex | 1,699,445 | 49,252 | 12,746 |
| Sex-dependent | 27,706 | 2,489 | 976 |
| Male-specific | 2,876 | 351 | 201 |
| Female-specific | 1,981 | 255 | 153 |
| Male-stratified | 865,986 | 31,384 | 9,507 |
| Female-stratified | 578,481 | 25,180 | 8,264 |

We found 49,252 CpG sites with a cross-sex mQTL and 2,489 CpG sites with sex-dependent mQTL. Of the CpG sites with sex-dependent mQTL, we found 351 CpG sites with a male-specific mQTL, and 255 CpG sites with a female-specific mQTL. Of the 351 CpG sites with a male-specific mQTL, 185 (53%) also had a cross-sex mQTL. Of the 255 CpG sites with a female-specific mQTL, 153 (60%) also had a cross-sex mQTL. 75 CpG sites had both male- and female-specific mQTL effects (see Figure 1 for an example of these types of mQTL effects), of which 74 (99%) also had a cross-sex mQTL (see Figures S1A-D for overlap between sets and for a comparison with male- and female-stratified mQTL).

Across chromosomes, we observed that the proportion of CpG sites with a cross-sex vs. sex-dependent mQTL differed for 10 of 23 chromosomes (Table S2, Figure 2A, chromosomes with a proportion test^25–27^ adjusted p < 0.05 marked with an asterisk). The proportion of male- vs. female-specific mQTL was more consistent across chromosomes (Figure 2B, for male- and female-stratified differences see Tables S2).

**Figure 2.**
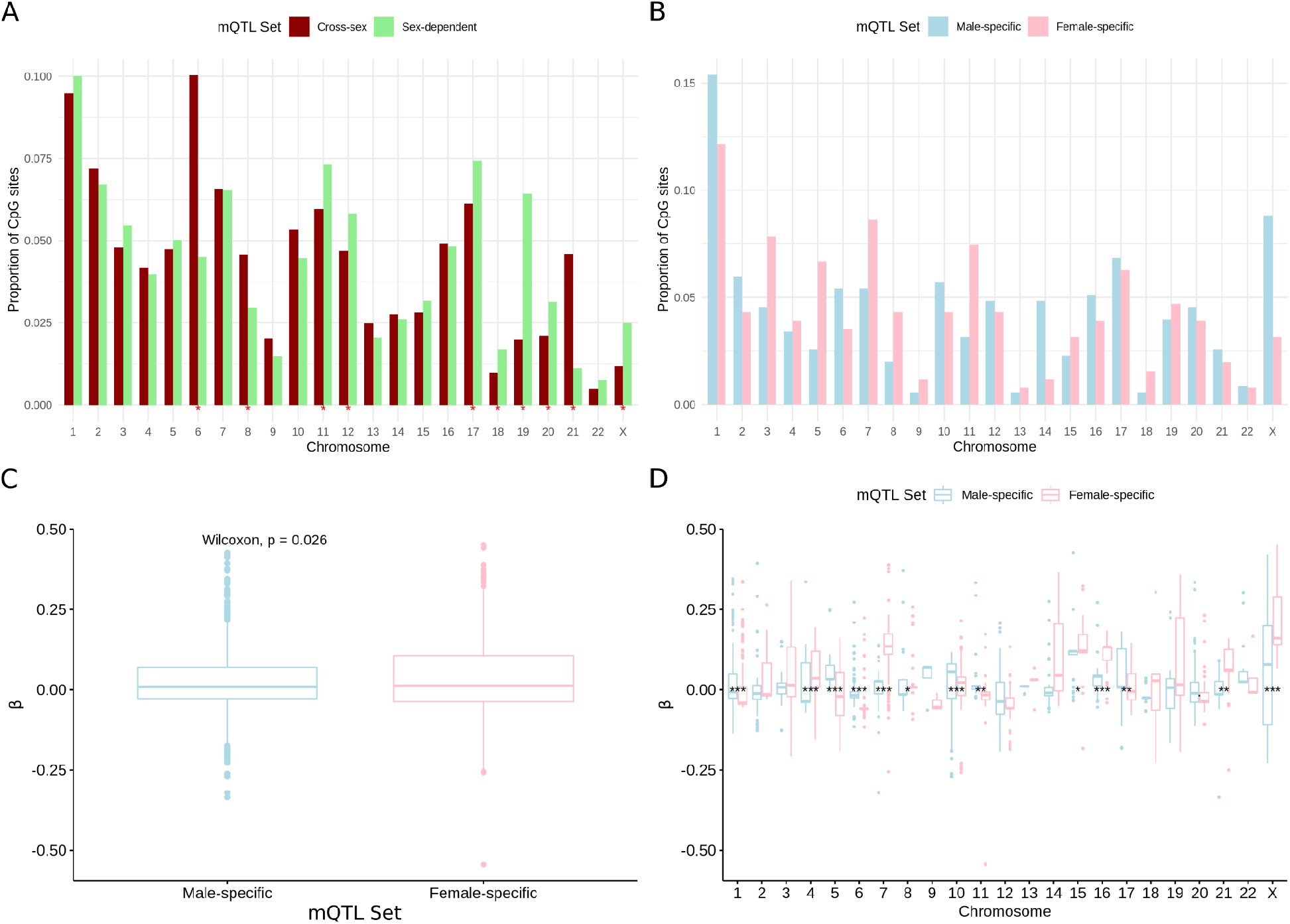
mQTL detected across chromosomes. (A) The proportion CpG sites with at least one cross-sex or sex-dependent mQTL by chromosome. Chromosomes with a significant difference in the proportion of cross-sex vs. sex-dependent mQTL are marked with an asterisk (FDR < 0.05, proportion test). (B) The proportion of CpG sites with at least one male- or female-specific mQTL called per chromosome. No chromosomes in this analysis had a significant difference in proportion of male- vs. female-specific CpG sites with at least one mQTL (i.e., meeting a FDR < 0.05, proportion test). (C) The difference in mean effect size in male- vs. female-specific mQTL across all chromosomes and (D) separately by chromosomes, with Wilcoxon rank-sum test p values after Bonferroni-Holm correction denoted as follows: .: p <=0.1; *: p <= 0.05, **: p <= 0.01;***: p <= 0.001.

We further investigated the difference in male- vs. female-specific beta values (the strength of SNP-CpG associations). Across adult tissues in GTEx V8 with sex-dependent eQTL, the majority of associations were in the same direction in both males and females, but had differences in effect size.^18^ We found a similar pattern. In our meta-analysis, the average effect in male-specific mQTL was smaller than the effect observed in female-specific mQTL (mean beta 0.021 in males, 0.035 in females, Wilcoxon rank-sum test two-sided p < 0.026, Figure 2C, Table S3). This difference was larger when we consider the absolute value of the effect size (mean absolute beta 0.067 in males vs. 0.079 in females, Wilcoxon rank-sum test two-sided p < 2.3e-11, Figure S1F, Table S3). This effect size difference did not appear to be driven solely by differences in mQTL effect sizes on the X chromosome: female-specific mQTL had a larger mean signed effect size than male-specific mQTL in 6 autosomes, and a larger mean absolute effect size in 9 autosomes (Wilcoxon rank-sum test one-sided, Bonferroni-Holm corrected p < 0.05, Figure 2D, Table S3). Effects were highly correlated between male- and female-specific mQTL (Spearman’s ρ = 0.87, Figure S1G). Thus, despite representing mQTL specific to one sex, male- and female-specific mQTL tended to have the same direction of effect.

### Placental mQTL are enriched in regions controlling gene expression and primarily occur at CpG sites with intermediate levels of DNAm

In order to establish whether sex-dependent mQTL occur in distinct genomic regions compared to cross-sex mQTL, we applied GARFIELD^28^ to the minimum mQTL p value for each SNP (STAR Methods). Briefly, GARFIELD tests whether a given set of SNPs associated with a particular phenotype are enriched in a set of genomic regions defined from functional experiments from the ENCODE Project Consortium^29–31^ and the NIH Roadmap Epigenomics Consortium.^32^ Importantly, GARFIELD accounts for both linkage disequilibrium (LD) between SNPs and redundancy in annotations: it will not overestimate enrichment due to many correlated mQTL in the same genomic region, and it will penalize annotations with similar enrichment across all mQTL in a set.

As expected, the enrichment of each mQTL set in each annotation was typically larger for sets with a larger number of mQTL, with cross-sex having the most mQTL, followed by male-stratified mQTL, female-stratified mQTL, and then sex-dependent mQTL (Table 1). Male- and female-stratified mQTL were most enriched within the same annotations as in cross-sex mQTL in the same order, albeit with smaller estimates. Male- and female-specific mQTL were excluded here, as GARFIELD uses only a single p value for each SNP and therefore could not handle the main and interaction term p values that define the male- and female-specific analyses. Across ChromHMM annotations in human cell lines (Figure 3A)^30,33^ we observed that cross-sex placental mQTL were more enriched in transcription start sites (TSS) than in other chromatin states (TSS odds ratio (OR) of 2.68 vs. a mean OR=1.60 in the remaining selected states), which was not observed for sex-dependent mQTL (TSS OR=1.22 vs. a mean OR=1.47 across other annotations). Within individual histone modifications averaged across experiments from ENCODE,^28,29^ we found that cross-sex mQTL were most enriched in regions with H3K9 acetylation (mean OR=2.50), followed by H3K4 trimethylation (mean OR=2.41), both of which are indicative of active gene promoters (Figure 3B).^34^ For sex-dependent mQTL, the highest enrichment was in H4K20 monomethylation (OR=1.69), which is associated with transcription activation.^35^ Overall, enrichment tracked strongly with the size of each mQTL set, and results suggested minor, but potentially meaningful, differences in the gene regulatory function of sex-dependent vs. cross-sex mQTL.

**Figure 3.**
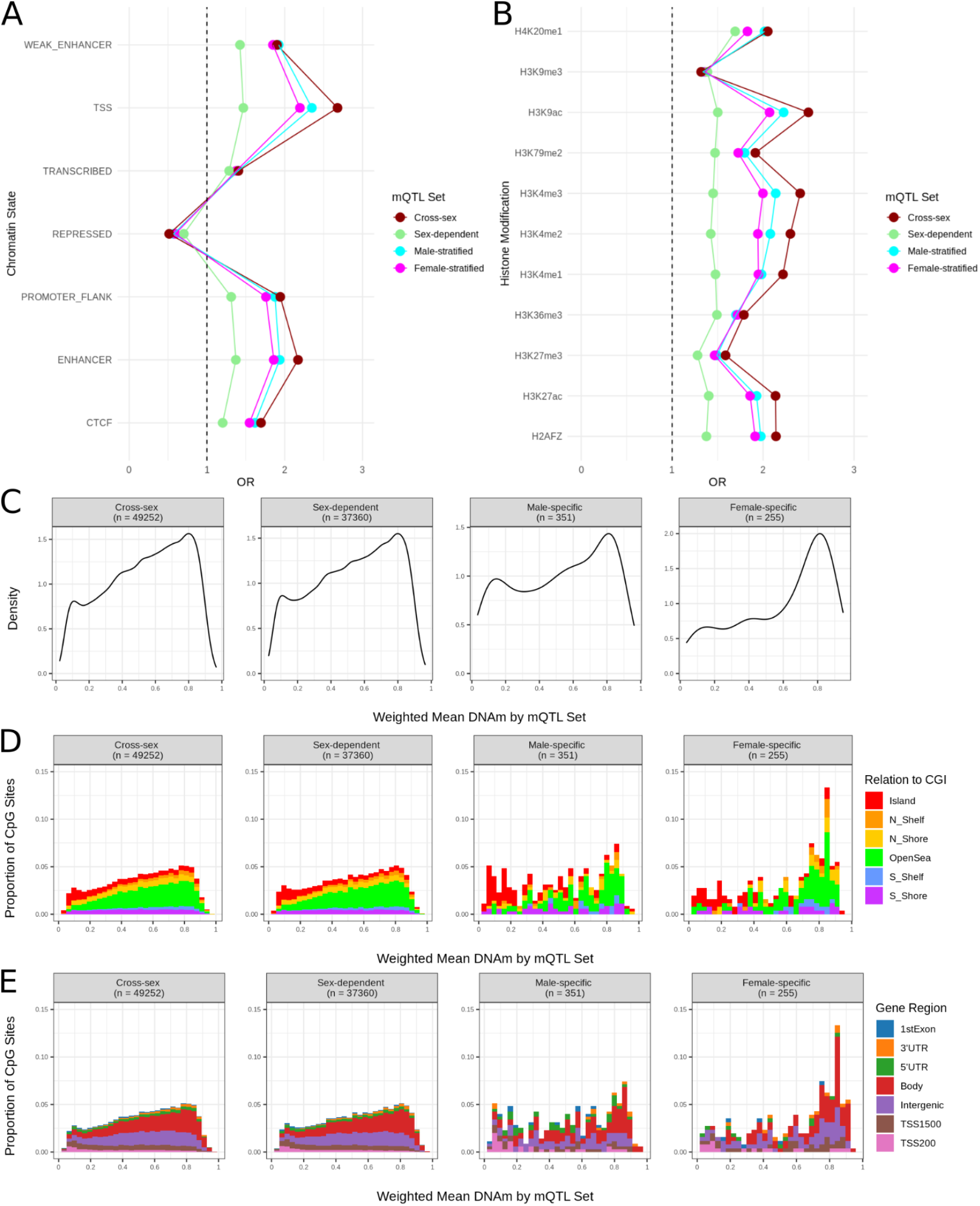
Functional role of placental mQTL. Enrichment of mQTL sets in chromatin states averaged across human stem cell lines (A) and in histone modifications averaged across tissues in ENCODE (B). (C) Density distribution of weighted mean DNAm, stratified by mQTL set. Proportion of CpG sites by weighted mean DNAm values, stratified by mQTL set, and visualized according to position relative to CpG island (D) and gene region (E).

Next, we investigated the weighted-mean DNAm of CpG sites with at least one cross-sex, sex-dependent, male-specific, or female-specific mQTL. Both cross-sex and sex-dependent mQTL were primarily associated with CpG sites with intermediate levels of DNAm. 72% of cross-sex and 71% of sex-dependent CpG sites having a weighed mean beta between 0.2 and 0.8 (Figure 3C). These proportions are much larger than what is observed in other human primary cell types (2% of sites with intermediate DNAm)^36^ and is not surprising given that up to 40% of CpG sites in the placenta are intermediately methylated.^37,38^ CpG sites corresponding to male- and female-specific mQTL were also primarily intermediately methylated (60% and 55%, respectively), but 31% of female-specific CpG sites had a weighted mean beta>0.8, compared to 21% of male-specific sites, and 16% of cross-sex and sex-dependent sites.

We next investigated how the level of DNAm of CpG sites with mQTL were spread across regions related to gene regulation. Specifically, we looked at the weighted-mean DNAm of these CpG sites relative to CpG islands (CGIs) and gene regions, as mapped Illumina’s HumanMethylation450k array annotation (STAR Methods). CGIs are where the majority of DNAm occurs, and the location of a given CpG site relative to a CGI is related to its biological function. Broadly speaking, whether DNAm occurs within isolation or in close proximity to other CpG sites, or upstream or downstream (north or south) of regulatory element containing a CGI, will change hypotheses regarding how and where that CpG site affects transcription.^39^ For example, high DNAm in CpG shores (within 2kb of a CpG island) and shelves (within 2-4 kb of a CpG island) is associated with higher nearby gene expression.^40^ The position of mQTL, and their associated CpGs, relative to CGIs and genomic regions can inform whether a given genetic variant is likely to affect DNA methylation, and gene expression over a broader stretch of DNA.^8^

We observed a similar distribution of DNAm levels per CGI region across our four mQTL sets of interest. The majority of highly methylated (beta > 0.8) CpG sites were in open sea regions (51% of cross-sex, 52% of sex-dependent, 57% of male-specific and 52% of female-specific), followed by regions within shores or shelves (36% of cross-sex, 36% of sex-dependent, 35% of male-specific, and 38% of female-specific), and the remaining CpG sites within CGIs themselves (13% of cross-sex, 12% of sex-dependent, 10% of male-specific, and 10% of female-specific) (Figure 3D). Relative to gene regions, we also observed similar patterns of DNAm levels across all four mQTL sets. Highly methylated CpG sites were primarily located in gene bodies (51% of cross-sex, 51% of sex-dependent, 56% of male-specific, and 53% of female-specific), which typically indicates active transcription of those genes. Intermediately methylated CpG sites were evenly split between gene bodies and intergenic regions (37% and 33% for both cross-sex mQTL and sex-dependent mQTL, 37% and 30% for male-specific mQTL, and 38% and 30% for female-specific mQTL respectively). Finally, lowly methylated CpG sites were primarily within 1500bp of the TSS of genes (39% in cross-sex, 39% in sex-dependent, 41% of male-specific, and 43% of female-specific), indicating active transcription (Figure 3E).

### Sex-dependent and cross-sex mQTL show similar patterns of tissue specificity

To assess the tissue specificity of the placental mQTL we identified, we first calculated the replication (π_1_) of placental mQTL across mQTL in two other prenatal tissues, umbilical cord blood and fetal brain, identified in independent datasets (Figure 4A; STAR Methods).^41,42^ We observed a relatively large proportion of placental mQTL overlapping cord blood mQTL (π_1_=0.76) and and even larger proportion of placental mQTL overlapping fetal brain mQTL (π_1_=0.84). Effect sizes of placental mQTL correlated poorly with those in cord blood (Spearman’s ρ=-0.31) but largely shared the same direction of effect in fetal brain (Spearman’s ρ=0.65), which suggests a higher degree of similarity between mQTL in placenta vs. in fetal brain.

**Figure 4.**
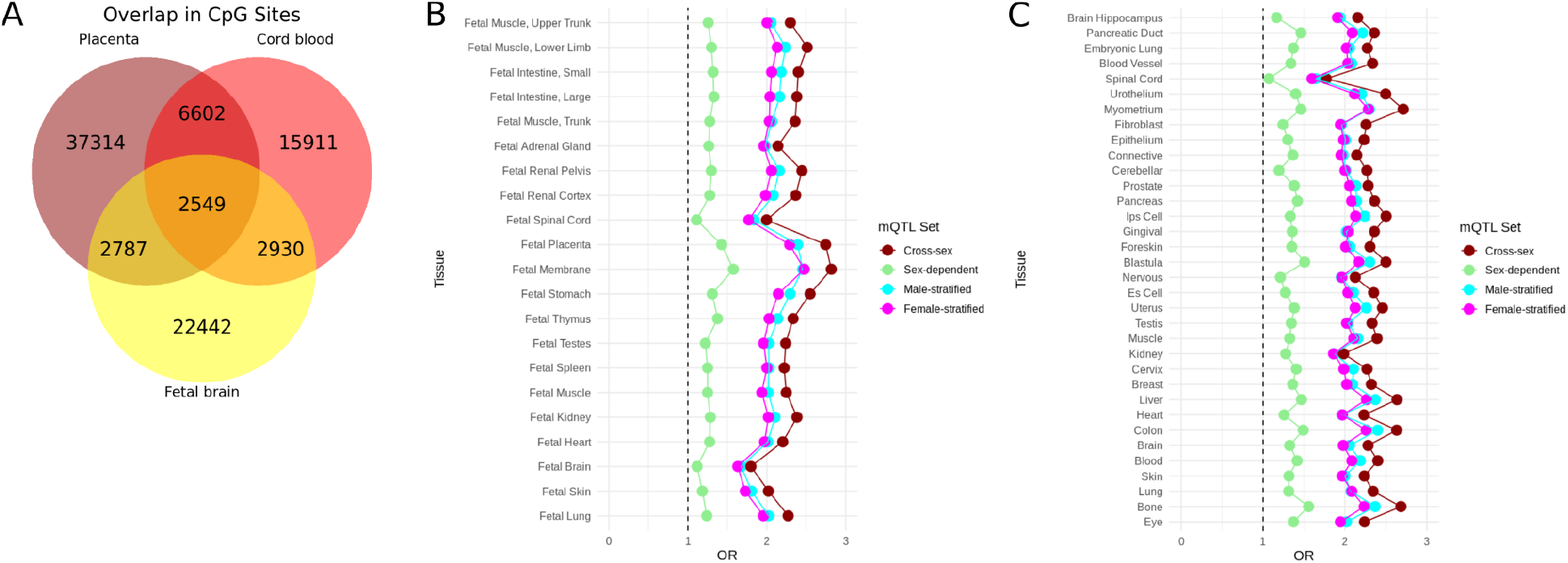
Enrichment of placental mQTL across tissues. (A) Overlap in CpG sites with at least one mQTL across prenatal tissues: umbilical cord blood from the ARIES study and whole fetal brain tissue from the HDBR study. Enrichment of placental mQTL in DNAse hypersensitivity sites in fetal tissues from the NIH Roadmap Epigenomics Consortium (B) and adult tissues from the ENCODE Consortium and the NIH Roadmap Epigenomics Consortium (C). Estimates are averaged over samples from the same tissue and stem cell lines.

Next, we quantified the enrichment of our mQTL sets in DNAse1 hypersensitivity (DHS) sites in fetal tissues, as well as in adult tissues and cell lines studied in the ENCODE Project Consortium and the NIH Roadmap Epigenomics Consortium,^29–32^ using GARFIELD. Enrichment was highest in fetal membrane for both cross-sex mQTL (OR=2.82) and sex-dependent mQTL (OR=1.58), followed by fetal placenta (Figure 4B, OR=2.74 and OR=1.43 in cross-sex vs. sex-dependent mQTL respectively). For adult tissues and cell lines (Figure 4C), cross-sex mQTL were most enriched in myometrium, which is tissue from the uterine wall (OR=2.71), followed by bone (OR=2.68), liver (OR=2.64), and colon (OR=2.64). Slight differences were found in sex-dependent mQTL, with enrichment being highest in bone (OR=1.56), blastula (an early stage of embryonic development, OR=1.51), colon (OR=1.49) and liver (OR=1.47). Overall, these results suggest that cross-sex and sex-dependent mQTL are enriched in regions that are active in the same sets of tissues.

### Male- and female-specific placental mQTL are more enriched for heritability of immune-related and growth-related traits than are cross-sex placental mQTL

In this study, we were particularly interested in how placental mQTL, including those specific to males or females, contributed to the genetic risk of childhood traits and conditions. We included GWAS of 18 childhood traits (Table SI; Figure 5A),^43,44^ as well as a GWAS of maternal pre-eclampsia (the fetal genetic effect on maternal pre-eclampsia risk),^45^ which is a placentally-mediated condition. We then used S-LDSC to estimate the proportion of SNP-heritability (h^2^_SNP_) of these 19 complex traits that was explained by our placental mQTL sets. We grouped childhood traits (excluding maternal pre-eclampsia) into three GWAS categories: neuropsychiatric (N=7), immune-related (N=3), and growth-related (N=8), and meta-analyzed enrichment results across traits within each category. We found that placental mQTL were not enriched for h^2^_SNP_ of any neuropsychiatric traits, either individually, or when all 6 traits were meta-analyzed (see Table SII for trait-level S-LDSC results, and Table S4 for meta-analyzed results). Conversely, we found an enrichment (cross-trait FDR < 0.05) of cross-sex placental mQTL in type 1 diabetes, birth weight, and pubertal growth start (Figure 5B). We also observed an enrichment for h^2^_SNP_ of child onset asthma, birth length, and late pubertal growth at a within-trait FDR < 0.05.

**Figure 5.**
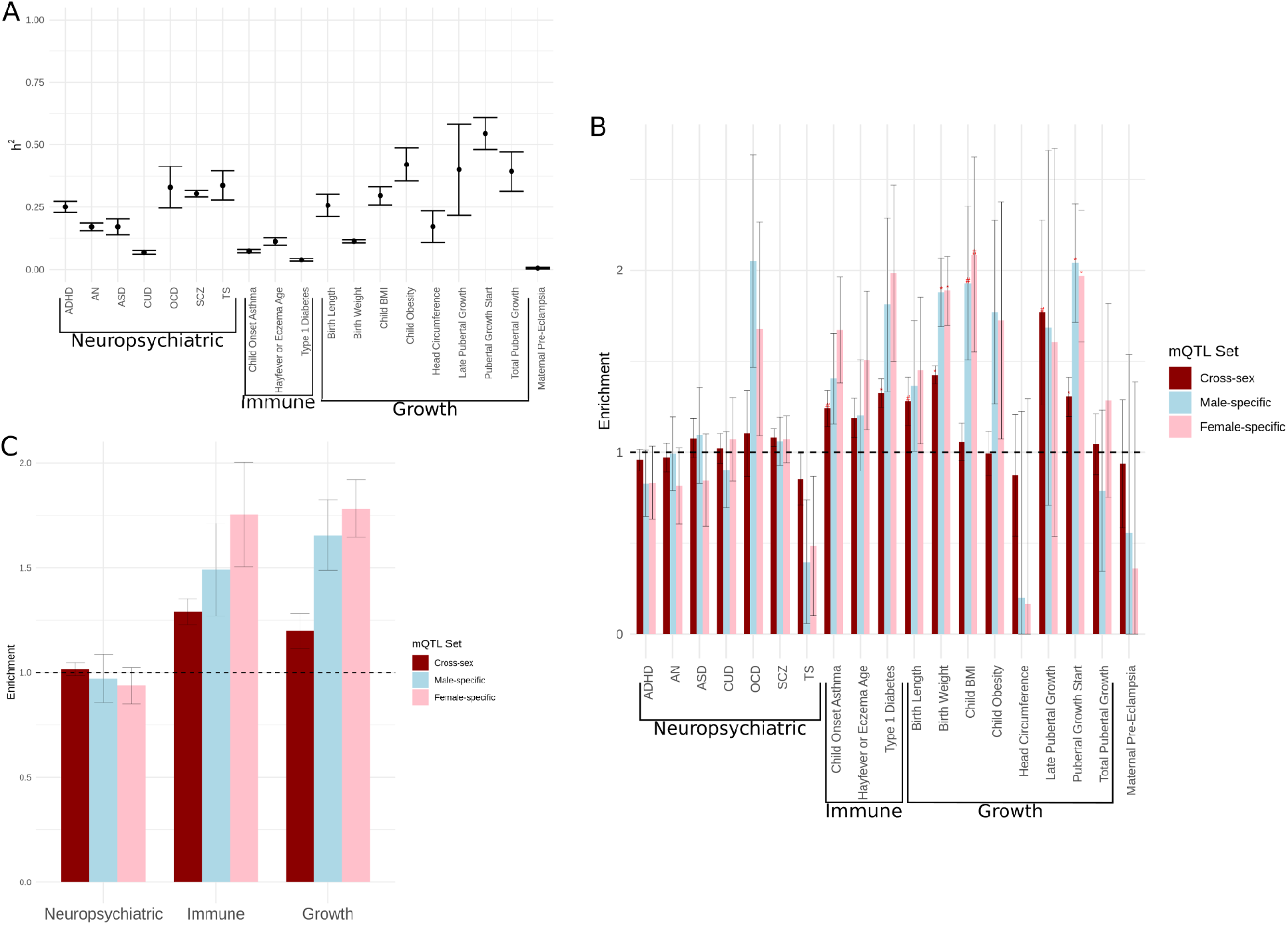
Placental mQTL enrichment in 19 complex traits. (A) SNP heritability (h^2^_SNP_) for each trait estimated by LD score regression. (B) Enrichment of placental mQTL sets for h^2^_SNP_ of each trait, accounting for 97 standard baseline regulatory effects. Enrichments with a within-trait FDR < 0.05 are marked with #, whereas enrichments significant at an FDR < 0.05, accounting for each mQTL annotation and GWAS assessed, are marked with an asterisk. (C) Meta-analyzed enrichment estimates across traits in neuropsychiatric, immune-, and growth-related GWAS categories.

Male- and female-specific mQTL, were enriched for h^2^_SNP_ of birth weight and pubertal growth start (FDR < 0.05), as well as of childhood BMI (within-trait FDR <0.05). In meta-analysis, female-specific mQTL were more enriched than cross-sex mQTL for h^2^_SNP_ of immune-related traits (1.75, SE=0.249 for female-specific mQTL vs. 1.29, SE=0.06 for cross-sex mQTL) (Figure 5C). Additionally, both male- and female-specific mQTL were more enriched than cross-sex mQTL for h^2^_SNP_ of growth-related traits (1.66, SE = 0.167 for male-specific mQTL, 1.78, SE=0.137 for female-specific mQTL, and 1.20, SE= 0.08 for cross-sex mQTL).

We also examined sex-stratified h^2^_SNP_ estimates and the proportion of sex-stratified h^2^_SNP_ explained by our mQTL sets. Since the sex-stratified GWAS sample sizes were smaller, fewer traits passed our quality control checks (STAR Methods): four neuropsychiatric traits (two with both sexes), one immune-related traits (with both sexes), and three growth-related traits (two with both sexes) (Figure S2A). Male- and female-specific mQTL were enriched for the sex-stratified h^2^_SNP_ of pubertal growth start at an FDR < 0.05 (Figure S2B, for meta-analyzed enrichment estimates see Figure S2C).

Overall, these results show that placental mQTL show larger enrichments in immune- and growth-related traits, with notably high enrichment in anthropometric traits like birth weight. Moreover, male- and female-specific mQTL all showed a larger enrichment for immune- and growth-related traits with significant enrichments than either cross-sex, male- or female-stratified mQTL, which highlights the importance of considering sex-dependent mQTL in measuring the enrichment of h^2^_SNP_ for these traits.

### Placental mQTL colocalize primarily with growth and immune related traits, with additional CpG sites colocalizing with male- and female-specific placental mQTL

We subsequently applied colocalization analyses using *coloc* (STAR Methods)^46^ to assess whether any of the mQTL we identified were also likely to be associated with the GWAS loci of 18 childhood traits and maternal pre-eclampsia. We conducted this analysis for cross-sex and sex-stratified mQTL, for GWAS loci within a 150kb window centered on each CpG site. We restricted this analysis to sex-stratified and not sex-specific mQTL as colocalization was conducted on mQTL p values, and the sex-specific mQTL are defined by their interaction with sex. We found a considerable number of colocalized CpG sites in several of the childhood traits with moderate enrichment in S-LDSC, such as child onset asthma, type 1 diabetes, and birth weight (Figure 6A, Table SIII). We also observed a high number of colocalized sites for SCZ, which notably has a large number of GWAS loci.^47^ Notably, loci from each of these four traits showed colocalization with male- and female-stratified mQTL that was not detected with cross-sex mQTL, and we label these male- and female-specific colocalized mQTL (see Figure 6B-G for examples of these distinctions).

**Figure 6.**
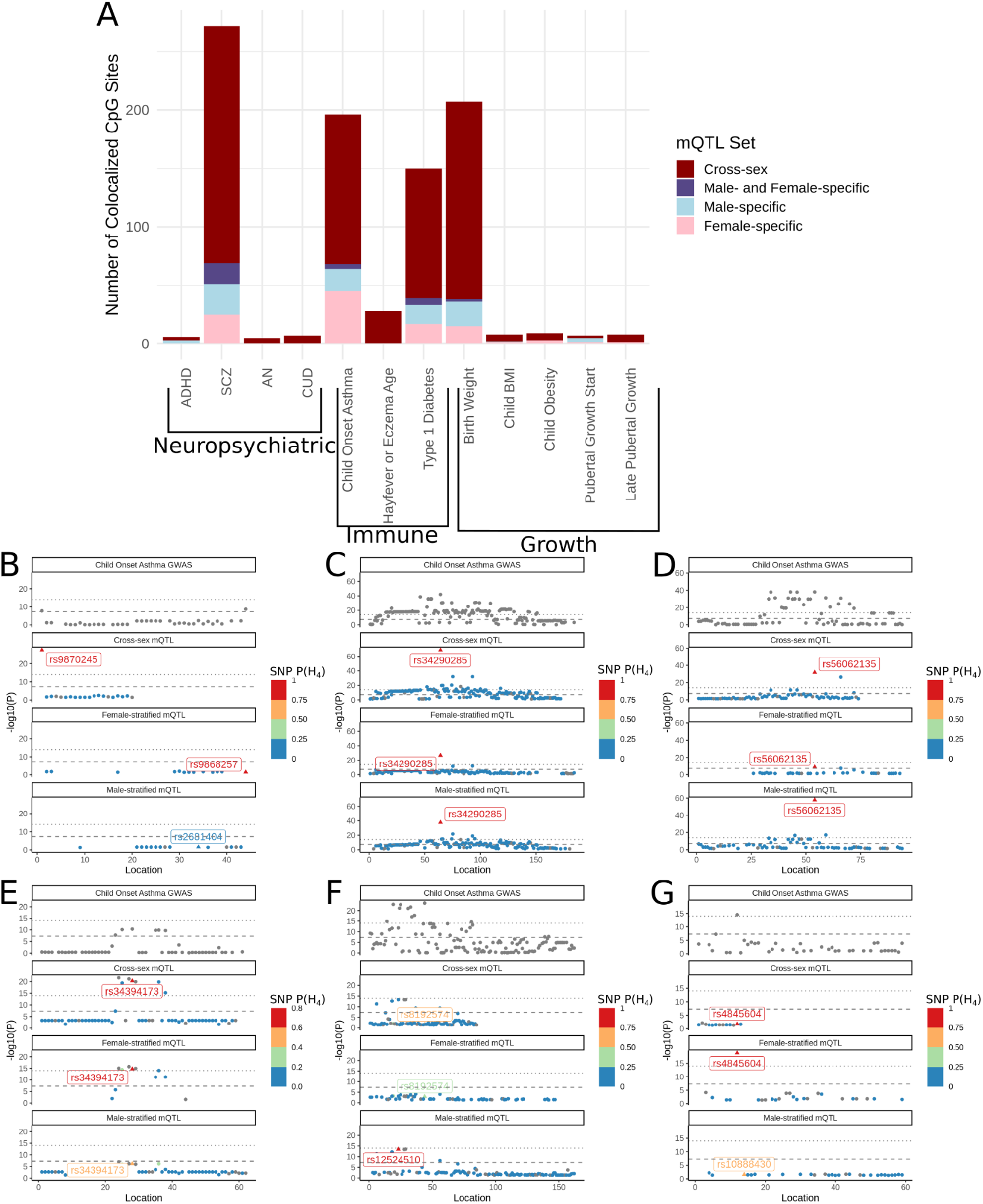
Colocalization of placental mQTL with GWAS loci of complex traits. A) The number of CpG sites with an mQTL that colocalized with at least one GWAS locus at a posterior probability (H_4_) > 0.9. We show examples of different colocalization scenarios that occurred when testing for colocalization of male- or female-stratified vs. cross-sex mQTL and loci from a GWAS of child onset asthma. In these plots (B-G), the location of each SNP is plotted from lowest to highest base-pair. We color by the per-SNP P(H_4_) for each CpG site with the highest overall P(H_4_) in different mQTL sets, and the SNP with the highest SNP P(H_4_) for each CpG site is labeled with a triangle and SNP identifier. We show colocalization in: (B) cross-sex mQTL only (cg16796354, in an intergenic region); (C) cross-sex, male- and female-stratified mQTL (cg02698622, within *RAPGEFL1*); (D) cross-sex and male-stratified mQTL (cg24032190, within *SMAD3*); (E) cross-sex and female-stratified mQTL (cg19063856, within *ILDR1*); (F) male-stratified mQTL only (cg16689962, within *D2HGDH*); and (G) female-stratified mQTL only (cg01717973, within *ILDR1*).

We used child onset asthma as our example to contrast the genes containing CpG sites colocalized with cross-sex vs. male-specific vs. female-specific mQTL. Sexual dimorphism in child onset asthma is known: with males having a higher prevalence of asthma up to age 13, and females having a higher prevalence of asthma in adulthood, often with greater severity of symptoms.^48^ Importantly, inasmuch as child onset asthma has a distinct genetic architecture from adult onset asthma (onset after age 19, genetic correlation, *r*_*g*_ = 0.67), genetic variants in the human leukocyte antigen (HLA) region, are frequently associated with both forms of the trait.^49,50^ The HLA region also contains numerous variants associated with allergic diseases.^51^

A total of 69 child onset asthma loci and colocalized with placental mQTL, of which 54 were cross-sex, five of which were male-specific, and 10 of which were female-specific. Of the 54 colocalizing cross-sex mQTL, 14 (26%) were annotated to genes from the HLA region (using Illumina’s HumanMethylation450k annotation). We mapped the remaining 40 cross-sex colocalized CpG sites (outside of the HLA region) to genes and then used Enrichr^52–54^ to identify enriched pathways at an adjusted p value < 0.05, and at a minimum overlap of 3 genes, across the three main genome ontology annotations. These were most enriched for the regulation of type I interferon gamma production, which is important for inflammation and also contains genetic variants with a negative vs. positive effect on asthma incidence in males vs. females respectively (adjusted p < 4.8e-4, GO:0032479, overlapping *TRAF3, IRF1, STAT6, POLR3H*, and *NFKB1*).^48,55^ In comparison, of the 10 colocalized female-specific CpG sites, 5 were in the HLA region and 3 mapped to genes of the major histocompatibility complex class III (*BAT1, BAT2*, and *BAT3*).^56^ Of the 5 colocalized male-specific CpG sites, 3 were in the HLA region, 1 mapped to the gene *NEU4*, which is related to metabolism, and one mapped to the gene *TSBP1* (*C6ORF10*), which encodes a protein most highly expressed in the testis.^57^

## Discussion

In this study, we conducted a meta-analysis of mQTL across two term placental studies (NICHD and RICHS), and a comprehensive sex-dependent mQTL analysis. We demonstrated that sex-dependent mQTL are located at partially distinct CpG sites from cross-sex mQTL. In contrast to blood mQTL, which are associated with mainly lowly or highly methylated CpG sites,^9^ placental mQTL tended to be associated with intermediately methylated CpG sites.^37^ Additionally, we showed that cross-sex mQTL were enriched for h^2^_SNP_ of immune- and growth-related traits and that male- and female-specific mQTL were more enriched than cross-sex mQTL across traits in these GWAS categories. Perhaps most importantly, we found several CpG sites with only male- or female-specific colocalization with GWAS loci. Thus, sex-specific mQTL appear to capture some underlying aspect of heritable trait risk that is not being captured by cross-sex placental mQTL alone.

Motivated by the enrichment of placental mQTL for the h^2^_SNP_ of immune-related traits, we homed in on asthma and mapped the colocalized asthma GWAS and mQTL to genes and pathways. Many of the colocalized loci were within the HLA region, which is frequently associated with child onset asthma and allergic diseases as well as with asthma hospitalizations in adults.^49–51,58^ Epidemiological studies have associated abnormal placental morphology, low birth length, and low birth weight to asthma risk.^59^ Our study suggests that genetic risk for child onset asthma is meaningfully enriched for cross-sex and male- and female-specific mQTL, and we provide these associations as a resource for potentially understanding the immediate biological consequences of child onset asthma loci.

Past eQTL ^12,15,16^ and integrative mol QTL analyses in placenta,^14,60^ have identified genetically regulated placental gene expression that associates with several traits, including child BMI, birth weight,^16,60^ asthma, and type 2 diabetes.^14^ All of these traits have evidence of genetic sexual dimorphism, either having a *r*_*g*_ lower than 1, or a difference in male vs. female effect size at risk loci.^48,61,62^ Likewise, the prevalence and manifestation of these traits differs between males and females throughout lifecourse.^48,63,64^ Importantly, the placenta is a sexually dimorphic tissue: we and others have found as many as 2,745 CpG sites (annotated to 582 genes) associated with placental sex, which annotated to genes that are primarily related to immune function and growth-factor signalling.^19,20^ Given these molecular sex differences and their potential role in mediating complex traits, sex-dependent effects should remain a focus of future studies of placenta.

There are several limitations to this study. First, our understanding of the tissue-specificity of placental mQTL effects is limited, as efforts are still underway to characterize mQTL across tissues.^65^ However, based on existing datasets, we showed that cross-sex placental mQTL quite similar to mQTL in other prenatal tissues (π_1_=0.76 in umbilical cord blood and π_1_=0.84 fetal brain, see Results). By this metric, placental mQTL are more similar to prenatal tissue mQTL than placental eQTL are to eQTL in 44 adult tissues from GTEx (ranging from π_1_=0.32 in cerebral hemisphere to π_1_=0.69 in fibroblasts; previously computed by others).^12^ Second, sex-dependent mQTL are less numerous than cross-sex mQTL due to the increased power required to detect interaction effects.^66^ As a result, in all of our experiments comparing sex-dependent mQTL (and by extension, male- and female-specific mQTL) to cross-sex mQTL, sex-dependent mQTL had larger standard errors, which hampered statistical inference and ultimately hampered our ability to discuss small differences in heritability captured by different mQTL sets. In particular, this made it difficult to draw conclusions from the τ^*^ metric, which can be more sensitive than the enrichment metric to small differences in mQTL annotations in single traits.^23^ We computed τ^*^ here to make it easier for readers to detect large differences between S-LDSC enrichment of placenta mQTL vs. mQTL from other tissues. As we accrue larger sample sizes in molecular studies of placenta and other tissues, we will be better equipped to identify robust differences in the contribution of male- and female-specific mQTL to the h^2^_SNP_ of complex traits. Third, we were limited in our ability to test male- and female-specific mQTL for enrichment in sex-stratified GWAS corresponding to their respective sex. Although sex-stratified GWAS results are increasingly being shared,^67^ these GWAS are underpowered. Only the GWAS of ADHD, ASD, SCZ, hayfever or eczema age of onset, pubertal growth start, and total pubertal growth were sufficiently powered to meet our criteria for inclusion in the enrichment analyses in either sex. For these traits, we found higher enrichments of male- and female-specific mQTL as compared to enrichments found for sex-pooled GWAS, which emphasizes the need for large sex-stratified GWAS.

Lastly, LDSC methods have mainly been validated in reference panels matching the genetic ancestry of GWAS samples.^21–23^ Here we made use of the 1000 Genomes European reference panel and assessed effects in European GWAS summary statistics, thus ignoring effects that may arise in other populations. Our discovery and replication samples are ancestrally diverse, which means that the mQTL we identified are relevant beyond European populations. Nonetheless, mixed ancestry has been shown to impact the colocalization of eQTL and GWAS signal.^68^Therefore, our findings bear repeating using GWAS of ancestrally diverse populations. As our understanding of molecular traits and genetic associations in people of diverse ancestries grows, we will develop a better view of the biological mechanisms that underlie complex traits and conditions.

Overall, this study demonstrates that the genetic regulation of placental DNAm is partially sex-dependent. Sex-dependent placental mQTL can occur at distinct functional genomic regions from cross-sex placental mQTL, suggesting they have a distinct role in gene regulation. Placental mQTL explain a significant proportion of the h^2^_SNP_ across conditions related to immune function and growth, and both male- and female-specific mQTL are more enriched than cross-sex mQTL across these conditions, despite being far fewer in number. Trends we observed in the enrichment of placental mQTL translated to higher instances of colocalization between mQTL and GWAS loci from childhood onset traits and conditions, and using male- and female-specific colocalization allowed us to detect 216 CpG sites (annotated to 98 genes) that otherwise did not show sufficient evidence of colocalization with cross-sex mQTL. Taken together, our findings demonstrate that careful consideration of sex in mQTL analyses has the potential to provide additional information about the basis of complex traits, particularly when the tissue, molecular features, and traits queried are sexually dimorphic.

## Supporting information

Supplementary Figures and Small Supplementary Tables

Table SI. Information and accession for GWAS considered in this study

Table SII. Stratified LD score regression results for all mQTL sets

Table SIII. Counts of CpG sites and targeted genes colocalized with GWAS loci across cross-sex, male- and female-stratified placental mQTL

## Data Availability

All data produced are available online at https://osf.io/9r4wf/ All primary data referenced in this study are available upon database of Genotypes and Phenotypes (dbGaP) access request or otherwise available online:
Rhode Island Child Health Study: https://www.ncbi.nlm.nih.gov/projects/gap/cgi-bin/study.cgi?study_id=phs001586.v1.p1
https://www.ncbi.nlm.nih.gov/geo/query/acc.cgi?acc=GSE75248
Eunice Kennedy Schriver National Institute of Child Health and Human Development (NICHD):
https://www.ncbi.nlm.nih.gov/projects/gap/cgi-bin/study.cgi?study_id=phs001717.v1.p1

https://osf.io/9r4wf/

https://www.ncbi.nlm.nih.gov/projects/gap/cgi-bin/study.cgi?study_id=phs001586.v1.p1

https://www.ncbi.nlm.nih.gov/geo/query/acc.cgi?acc=GSE75248

https://www.ncbi.nlm.nih.gov/projects/gap/cgi-bin/study.cgi?study_id=phs001717.v1.p1

## Acknowledgements

This project was supported by a British Columbia Children’s Hospital Research Institute Catalyst Grant. JKD is supported by a NARSAD Young Investigator Grant from the Brain & Behavior Research Foundation, and is a Michael Smith Health Research BC Scholar.

phs001586 : Rhode Island Child Health Study (RICHS): This work was supported by the National Institutes of Health [NIH-NIMH R01MH094609, NIH-NIEHS R01ES022223, NIH-NIEHS PO1ES022832, NIH-NIEHS R24ES028507, NIH-NIEHS R21ES028226, and NIH-NIEHS R01ES025145]. phs001717: This research was supported by the Intramural Research Program of the Eunice Kennedy Shriver National Institute of Child Health and Human Development, National Institutes of Health (Contract Numbers:

HHSN275200800013C;HHSN27500006;HHSN275200800003I;HHSN275200800014C;HHSN2 75200800012C;HHSN275200800028C; HHSN275201000009C). We also want to thank the Genomics Shared Resource of Roswell Park Cancer Institute, supported by National Cancer Institute (NCI) grant P30CA016056.

This research was supported in part through computational resources and services provided by Advanced Research Computing at the University of British Columbia (https://doi.org/10.14288/SOCKEYE).

## Author Contributions

Conceptualization, W.C., J.K.D. and S.M.; Methodology, W.C.,Y.P, and J.K.D.; Software, W.C. A.I., J.D. V.Y.; Validation W.C.,A.I., J.D. V.Y.; Formal analysis, W.C.; Resources, J.K.D. and S.M.; Data Curation, W.C, C.M., and F.D.; Writing - Original Draft, W.C.; Writing - Review and Editing, J.K.D.; Visualization, W.C.; Supervision, J.K.D. and S.M.

## Declaration of Interests

The authors declare no competing interests.

## STAR Methods

### Data description

We obtained placental genotype and DNAm data from two different studies, both of which sampled fetal side placental tissue from pregnancies lasting >=37 weeks. Discovery sample: The National Institute of Child Health and Human Development (NICHD) study^12^ included 303 subjects (151 male, 152 female) of diverse reported maternal ethnicity (Table S1), the vast majority of which (96%) had no reported pre-eclampsia. Birth length was not publicly available for this study, and our discovery and replication samples could therefore not be compared to one another on this measure. All samples were genotyped using the Illumina HumanOmni2.5 Beadchip, and DNAm was measured using the Illumina HumanMethylation450k array (dbGaP accession phs001717.v1.p1).

Replication sample: The Rhode Island Child Health Study (RICHS)^15,17^ included 149 subjects (74 male and 75 female) of primarily non-hispanic white reported ethnicity. The samples were selected for being either large or small for gestational age (either > 90% or < 10% on the Fenton growth chart^69^). The majority of subjects (55.1%) are in the average range (i.e., in between 10% and 90%)(Table S1). No serious prenatal complications or congenital or chromosomal abnormalities were present in this sample. Samples were genotyped on the Illumina Expanded Multi-Ethnic Genotyping Array (Mega-EX) (dbGaP accession phs001586.v1.p1). DNAm was measured on the IlluminaHumanMethylation450k array (GEO accession GSE75248). DNAm and SNP genotype data were linked by a common ID file available upon request.

### Genotype data processing and quality control

Genotyping data from NICHD and RICHS were processed separately, and were subject to the same processing and quality control (QC) protocol, which was based on the RICOPILI pipeline.^70^ Using plink 1.9,^71^ we removed SNPs that were strand ambiguous, had a call rate < 0.05, or MAF < 0.01. We removed individuals with a mismatch between recorded and genotyped sex, SNP missingness > 0.02, and with excess heterozygosity (F_het_ > 0.2). SNPs on the X chromosome were kept, using the default 0/2 encoding in plink for SNPs on the X chromosome in males.

We used the GRAFpop algorithm^72^ on autosomal SNPs to assign individuals to one of 7 ancestry groups: EUR (European), AFR (African), AFR_AM (African American), LAT_AM_1 (Latin American 1), LAT_AM_2 (Latin American 2), EAS (East Asian), PAC (Asian Pacific Islander), and SAS (South Asian), which we used to help us decide the number of ancestry PCs to include in downstream analyses. Genetic ancestry assignments agreed well with maternal self-reported ethnicity (Table S5), and samples were filtered to ensure identity by descent did not exceed an |F_ST_| > 0.2 between samples within each ancestry group.

To define ancestry PCs, we first pruned SNPs based on LD in a pairwise manner using plink (running --indep-pairwise 200 100 0.2 twice), and then removed SNPs with an MAF < 0.05, and SNPs in highly recombinant regions (the HLA region, chr6:28,477,797-33,448,354 in GRCh37, and regions of long range LD).^73^ Ten ancestry PCs were computed from this set of SNPsusing FastPCA.^74^ From these 10, we ultimately selected the first five to include in all downstream analyses on the basis of their ability to separate distinct populations of subjects (defined by self-reported race or GRAF-pop estimated ancestry) in each study (Figures S3A-D).

Before imputing SNP genotypes, we removed variants that were associated with genotype batch (Bonferroni corrected p < 0.05) and variants with a Hardy-Weinberg equilibrium p < 1e-10. Variants were then aligned and imputed to the cosmopolitan 1000 genomes phase 3 reference panel^75^ using the Michigan imputation server.^76^ Variants were filtered to have an imputation quality R2 > 0.3 and an ancestry-specific MAF within 0.1 of their corresponding 1000 genomes phase 3 group. Last, any remaining imputed SNPs associated with genotype batch were removed. The NICHD study had 275 samples with 14,110,641 SNPs remaining for analysis, whereas RICHS had 136 samples with 6,139,984 SNPs remaining for analysis.

### DNA methylation array processing and quality control

Illumina HumanMethylation450k array data from the NICHD study and RICHS were processed separately using the bioconductor program *minfi*.^77^ Data from RICHS were first normalized for dye bias (*preprocessNoob, minfi*^77^) since the data were provided in raw array IDAT form as opposed to called beta values.

First the pattern of methylation across all probes was checked to confirm that it followed a hemi-methylated pattern characteristic of the placenta.^38^ Next, we removed individuals with sex mismatches, i.e., individuals with sex chromosome DNAm values that did not cluster with samples in their reported sex (two samples total, neither of which were flagged as sex-mismatched during genotyping QC). We then removed probes with a detection p > 0.01, and probes that either failed or were missing in > 20% of samples. Next, we removed SNP probes (annotated by Illumina and identified by others),^78^ Y chromosome probes, and non-specific cross-hybridizing probes.^79^ Lastly, the data were quantile normalized using *preprocessQuantile* from the R package *minfi*.^77^ After implementing these QC steps, the NICHD and RICHS studies had 447,232 and 446,976 probes remaining for analysis.

### Mapping cross-sex, sex-dependent, male-stratified, and female-stratified mQTL

We conducted four mQTL analyses to detect four different cis-mQTL effects: a cross-sex analysis, an interaction analysis (in which the effect of SNP on DNAm differed by sex, as captured by a genotype by sex interaction term), an analysis of males only, and an analysis of females only (Figure 1). Each analysis was conducted separately in NICHD and RICHS.

We used matrixEQTL^80^ for associating imputed genotype dosages to DNAm beta values. Each SNP within 75 kb upstream or downstream of each CpG site was regressed onto DNAm, accounting for gestational age, sex, methylation array batch, 5 genetic ancestry PCs, and 9 DNAm PCs. As interaction effects (i.e., sex-dependent effects) are harder to detect, and require a mQTL effect, we elected to exclude trans-mQTL, or SNPs > 75kb from an associated CpG site, from this analysis.^66,81,82^ We chose the 75 kb window as the majority of cis-mQTL in other tissues have been shown to be contained within this region.^12,83^ The number of DNAm PCs to include was determined based on the number of mQTL declared on chromosome 21 at a Bonferroni corrected p < 0.05, varying the number of PCs included while keeping other covariates fixed and selecting the number of PCs for which the number of mQTL declared did not improve (Figures S3E,F).^84,85^

Since sex-dependent molQTL detection can be biased by sex differences in cell type proportions,^18^ we ensured that the top 10 DNAm PCs were correlated with placental cell type proportions (Figures S4A,B). Placental cell type proportions were estimated using the data available on cell-type specific methylation in placenta in the R package *PlaNET*,^86^ and the Houseman algorithm implemented in *minfi*.^77^ Reassuringly, we found that cell-type proportion was most correlated with the top 5 DNAm PCs in both studies. We also checked if there was a difference in estimated cell-type proportion between males and females in each study (Figures S4C,D). We observed a sex difference in average estimated stromal cell proportion in the NICHD study, which is accounted for by DNAm PCs.

Before meta-analyzing results from the NICHD and RICHS samples, we tested cross-sex and sex-dependent mQTL identified in NICHD for replication in RICHS using the π_1_ statistic,^87^ which is a better measure of comparability than the raw proportion of overlapping sites because it accounts for between-study differences in the number of tested SNP-CpG pairs, which can artificially inflate or deflate estimates of overlap. For the π_1_ analysis, we relied on cross-sex and sex-dependent mQTL called in each study at an FDR < 0.05 because calling mQTL at a Bonferroni corrected p < 0.05 resulted in unstable estimates of π_1_. We identified cross-sex mQTL with linear models testing for association between genotype and DNAm. Sex-dependent mQTL were those with a statistically significant genotype by sex interaction term. In the NICHD study, we found 275,806 CpGs with at least one mQTL and 60537 CpGs with at least one mQTL that interacted with sex. In RICHS, we found 129,361 CpGs with at least one mQTL and 19,530 CpGs with at least one mQTL that differed by sex. The π_1_ estimate for the cross-sex mQTL was 0.74, which is the proportion of mQTL in the NICHD study that have similar effects in the RICHS study (computed over 2,691,024 SNP-CpG pairs available in both studies). Sex-dependent mQTL showed modest replication (π_1_ = 0.28, computed over 80,363 SNP-CpG pairs available in both studies), which is in line with values previously reported for sex-dependent eQTL in GTEx tissues (π_1_ = 0.28 in breast tissue, π_1_ ranging from 0-0.12 in the remaining 43 GTEx V8 tissues analyzed by Oliva et al.).^18^

We then meta-analyzed across the NICHD and RICHS samples for each of the four analyses using MeCS software,^24^ which was designed to account for the highly correlated nature of molecular cis-QTL effects between independent studies. Within each analysis, we retained significant mQTL, or mQTL with a significant genotype by sex interaction effect in the interaction analysis, where significance was declared at a Bonferroni corrected p < 0.05. From these four analyses, we defined six sets of mQTL: (i) cross-sex; (ii) sex-dependent; (iii) male-stratified; (iv) female-stratified; (v) male-specific; and (vi) female-specific. The (v) male- and (vi) female-specific mQTL were those with significant stratified and interaction effects (i.e., the intersection of sex-dependent and male- or female-stratified mQTL). The reported effect size for each male- and female–specific mQTL was taken from male- and female-stratified mQTL. In counting mQTL, associated CpG sites, and the genes to which CpG sites were annotated, SNPs with a MAF < 0.05 were excluded in order to ensure mQTL were less likely to be a result of outliers in DNAm.

### Comparing cross-sex placental mQTL to other perinatal tissues

As a main focus of this study is outlining the developmental origins of complex traits, we compared placental mQTL to mQTL from other tissues collected before and after birth. We accessed summary statistics for umbilical cord blood mQTL from the accessible resource for integrative epigenomic studies (ARIES; N=711; available via http://www.mqtldb.org/), originally from the Avon longitudinal study of parents and children,^41^ and fetal brain mQTL summary statistics from the human developmental biology resource^88^ (HDBR; N=173; available via https://epigenetics.essex.ac.uk/mQTL/). The ARIES study was comprised of 51% male samples, and in mQTL mapping authors accounted for sex, 10 ancestry PCs, batch, and cell-type proportion estimates for white blood cell counts. mQTL in the ARIES study were called at a p < 1 * 10^−14^ for all pairwise associations between 8,074,398 imputed SNPs at an MAF > 0.05 and 395,625 CpG sites passing their QC (i.e., cis- and trans-mQTL effects were identified). This threshold is more stringent than our Bonferroni corrected alpha < 0.05 computed for associations computed in cis, which corresponds to a threshold of p < 3.77 * 10^−10^. Meanwhile, the HDBR study in homogenized fetal brain tissue consisted of 54% male samples ranging from 8-24.1 weeks post conception. When mapping mQTL, they accounted for sex, age, and 2 genotyping PCs. Additionally, they had a more stringent methylation QC and included only directly genotyped SNPs at MAF > 0.05, resulting in 430,304 SNPs and 314,554 CpG sites tested for all pairwise associations (i.e., cis- and trans-mQTL effects). mQTL were called at a Bonferroni-corrected threshold of p < 3.69 * 10^−13^. Notably, the HDBR study excluded the sex chromosomes from their mQTL analysis.

### GARFIELD analysis

We used GARFIELD^28^ to quantify the enrichment of mQTL in different genomic regions across tissues, accounting for linkage disequilibrium (LD) and redundant annotations (i.e., annotations with similar enrichment to one another across mQTL). We used the genomic regions from the ENCODE and NIH Roadmap Epigenomics projects, which were provided as defaults by the software developers.^29–32^ GARFIELD was run for cross-sex, sex-dependent, male-stratified, and female-stratified mQTL sets by mapping each mQTL to its minimum p value regardless of the probe with which it was associated. As male- and female-specific mQTL were defined on two sets of p values, we elected to exclude them for this analysis. We then ran GARFIELD considering only mQTL that were significant at a threshold of 1 * 10^−9^ (accounting for the roughly 100 million SNP-CpG pairs tested in cross-sex, sex-dependent, male-stratified, or female-stratified mQTL analysis). To simplify our results, rather than reporting the enrichment result from each regulatory mark measured for each individual cell line, we chose to report the mean odds ratio of a regulatory mark across all cell lines taken from a single tissue.

### Linkage disequilibrium score regression analyses

We estimated GWAS trait h^2^_SNP_ using linkage disequilibrium score regression (LDSC) software^21,22,89,90^ (https://github.com/bulik/ldsc). We then partitioned h^2^_SNP_ by placental mQTL sets using stratified LDSC (S-LDSC). S-LDSC is most powerful when the annotation (in this case, mQTL sets) cover at least 1% of the genome. To expand our definition of mQTL to fit this criterion, and to maximize enrichment in our mQTL sets, we applied a previously developed method.^23^ Briefly, for each CpG site with at least one mQTL at an FDR < 0.05, we fine-mapped meta-analyzed mQTL associations with a nominal p < 0.05 using CAVIAR.^91^ The output from CAVIAR is a set of SNPs with a 95% likelihood of containing the SNP causal to changes in DNAm (i.e., 95% credible set). As fine-mapping for interaction effects is difficult to interpret, we did not consider sex-dependent mQTL as its own category for this analysis. Instead for CpG sites with at least one male-stratified or male-specific mQTL, as well as female-stratified or female-specific mQTL, we ran fine-mapping with male- and female-stratified mQTL p values respectively. We chose not to use the maximum causal posterior probability (max CPP) measure for each SNP as the weighted sum of our male- and female-specific annotations was small, less than 50,000 compared to the over 1 million per annotation that we achieved when using the 95% credible set (i.e., assigning 95% credible set SNPs a weight of 1, Figures S4E,F).

Once we had defined the mQTL sets for use in S-LDSC, the stratified LD scores for our annotations (i.e., mQTL sets) were built using all subjects available in the 1000 Genomes Phase III EUR population, the publicly available baseline v2.2 LD scores (consisting of 97 annotations) provided by the software developers, and excluding SNPs within the MHC region of the human genome.^21,22^ The EUR population was chosen as opposed to a transancestral population since LDSC has only been evaluated on EUR and EAS samples with ancestrally matched summary statistics, with little documentation of the effects of admixture,^21–23,89^ and the majority of GWAS used in our analyses were in European populations (see Availability and criteria for GWAS summary statistics, below).

We report enrichment as the proportion of h^2^_SNP_ explained by the SNPs in each mQTL set, divided by the proportion of SNPs included in the mQTL set. We also quantify enrichment using τ^*^ (Table SI), which is defined previously as:^89^

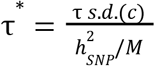

Where h^2^_SNP_ is the SNP heritability of the trait, s.d.(c) is the standard deviation of the annotation, τ is the coefficient of that annotation, and M is the number of variants used to compute h^2^_SNP_. Thus, tau can be interpreted as the standardized average contribution of SNPs in an annotation to the total h^2^_SNP_ of a trait. An advantage of τ^*^ over enrichment is that it quantifies effects that are specific to that annotation, i.e., after taking into account the overlap between the annotation of interest, and other annotations in the model. An elevated tau value suggests that enrichment is not explained by overlap with other annotations in the model. Negative τ^*^ values indicate that the annotation on its own reduces h^2^_SNP_, on average.

S-LDSC was run separately for cross-sex, male- and female-specific, and male- and female-stratified mQTL, accounting for the 97 baseline annotations. Following S-LDSC analysis in single traits, we meta-analyzed enrichment and τ^*^ estimates across trait groups using the *rmeta* package.^92^ Estimates from trait categories with sex-stratified GWAS summary statistics available were analyzed with mQTL annotations matching the sex of their component GWAS (GWAS labelled _female or _male in Table S4). τ^*^ and its standard error from meta-analysis were used to generate a normally distributed Z score for computing meta-analyzed p values. No such procedure is currently defined for computing significance from meta-analyzed S-LDSC enrichment values, so significance is not computed.

### Colocalization of placental mQTL and GWAS loci

We performed a colocalization analysis to characterize the extent to which the proportion of h^2^_SNP_ attributed to placental mQTL corresponded to shared genetic variants between mQTL and GWAS loci. We focused on all sets of SNPs within 75kb of CpG sites with a mQTL associated at a p < 5e-8, that were also associated with a GWAS trait at p < 5e-8, and we computed colocalization using the *coloc* R package.^46^ Notably, unlike our protocol in S-LDSC, we included the HLA region in colocalization analysis given that this analysis is not dependent on genome-wide patterns of LD, and the HLA region is of particular interest to the immune-related traits analyzed. All genes considered to be within the HLA region were selected from chr6:28,477,797-33,448,354 in genome build hg19, and were gathered from their corresponding table on the UCSC genome browser at http://genome.ucsc.edu.^93^ Colocalization was computed with cross-sex GWAS summary statistics. We defined colocalization having an overall P(H_4_) > 0.9 per CpG site, which corresponds to the likelihood of both the level of DNAm and the trait sharing a single causal genetic variant in that region.

In plotting these results (Figure 6A), we wanted to highlight the colocalizations that were found only through sex-stratified analyses. Therefore, all colocalizations identified in the cross-sex analysis were labeled as such, even if they were also identified in the male- and female-stratified analyses. A small number of colocalizations (N=30 CpG sites) were identified in both male- and female-stratified analyses (annotated to N=4 genes unique to this set), but not in cross-sex analyses. We label these “Male- and Female-specific” colocalizations. Finally, we plot colocalizations that were found in the male- and female-stratified analyses only, which we refer to as male-specific and female-specific colocalizations respectively.

### Availability and criteria for GWAS summary statistics

We define childhood onset traits and conditions as those which have a median age of onset < 25,^43^ which is roughly considered to be the end of brain maturation and adolescence.^44^ Summary statistics were formatted for all LD score analyses using the munge_sumstats.py script made available by the LDSC developers. This includes the following quality control measures: only biallelic SNPs are kept, strand ambiguous SNPs are excluded, duplicate SNPs are excluded, SNPs have an imputation INFO score > 0.9, a MAF > 0.01, 0 < p <= 1, SNPs with a number of samples < 90th quantile divided by 1.5 are excluded, and the median Z score of all SNPs are within 0.1 of its expected null value (0 for signed statistics, 1 for an odds ratio). This largely matches what has been done in previous publications dealing with sex-stratified summary statistics, with the exception of differences MAF threshold used across different studies^62,67^

Genome-wide summary statistics for all traits were downloaded in April 2021 and are publicly available (Table SI). Sex-stratified GWAS from the Psychiatric Genomics Consortium (PGC) were obtained from Martin et al, 2021.^67^ Summary statistics for preeclampsia were requested separately via an InterPreGen access request. In S-LDSC analyses, we imposed a filter of a GWAS h^2^_SNP_ z-score^21,22^ > 1.644854, corresponding to a h^2^_SNP_ estimate that was significant at a nominal p < 0.05.

## Large Supplemental Table Legends

**Table SI. Information and accession for GWAS analyzed in this study**..

**Table SII. Stratified LD score regression results for all mQTL sets and GWAS traits**.

**Table SIII. Counts of CpG sites and targeted genes colocalized with GWAS loci across cross-sex, male- and female-stratified placental mQTL**. Colocalized CpG sites were enumerated for each mQTL set considering overlap between mQTL sets.

## Notes

### Competing Interest Statement

The authors have declared no competing interest.

### Author Declarations

Data is available via an access request to the database of Genotypes and Phenotypes (dbGaP) or are otherwise publicly available online: Rhode Island Child Health Study: https://www.ncbi.nlm.nih.gov/projects/gap/cgi-bin/study.cgi?study_id=phs001586.v1.p1 https://www.ncbi.nlm.nih.gov/geo/query/acc.cgi?acc=GSE75248 Eunice Kennedy Schriver National Institute of Child Health and Human Development (NICHD): https://www.ncbi.nlm.nih.gov/projects/gap/cgi-bin/study.cgi?study_id=phs001717.v1.p1

### Summary of Updates

Mislabelled subfigures within text have been revised. Figure captions and supplementary figure captions have been revised. Modified figure 3 to include overlap with mQTL from different tissues; modified figure 5 to include LD score regression results; modified figure 6 to include examples of colocalization, thus removing LD score regression results from this figure. Edits have been made to grammar and punctuation throughout manuscript. References have been updated for main manuscript and Key Resources table

## References

1. King EA, Davis JW, Degner JF. Are drug targets with genetic support twice as likely to be approved? Revised estimates of the impact of genetic support for drug mechanisms on the probability of drug approval. PLoS Genet. 2019 Dec 12;15(12):e1008489.

2. Nelson MR, Tipney H, Painter JL, Shen J, Nicoletti P, Shen Y, et al. The support of human genetic evidence for approved drug indications. Nat Genet. 2015 Aug;47(8):856–60.

3. Maurano MT, Humbert R, Rynes E, Thurman RE, Haugen E, Wang H, et al. Systematic Localization of Common Disease-Associated Variation in Regulatory DNA. Science. 2012 Sep 7;337(6099):1190–5.

4. Schork AJ, Thompson WK, Pham P, Torkamani A, Roddey JC, Sullivan PF, et al. All SNPs Are Not Created Equal: Genome-Wide Association Studies Reveal a Consistent Pattern of Enrichment among Functionally Annotated SNPs. PLoS Genet. 2013 Apr 25;9(4):e1003449.

5. THE GTEX CONSORTIUM. The GTEx Consortium atlas of genetic regulatory effects across human tissues. Science. 2020 Sep 11;369(6509):1318–30.

6. Gamazon ER, Segrè AV, van de Bunt M, Wen X, Xi HS, Hormozdiari F, et al. Using an atlas of gene regulation across 44 human tissues to inform complex disease-and trait-associated variation. Nat Genet. 2018 Jul;50(7):956–67.

7. Umans BD, Battle A, Gilad Y. Where Are the Disease-Associated eQTLs? Trends in Genetics [Internet]. 2020 Sep 7 [cited 2020 Nov 18]; Available from: http://www.sciencedirect.com/science/article/pii/S0168952520302092

8. Do C, Shearer A, Suzuki M, Terry MB, Gelernter J, Greally JM, et al. Genetic–epigenetic interactions in cis: a major focus in the post-GWAS era. Genome Biol [Internet]. 2017 Jun 19 [cited 2020 Nov 23];18. Available from: https://www.ncbi.nlm.nih.gov/pmc/articles/PMC5477265/

9. Min JL, Hemani G, Hannon E, Dekkers KF, Castillo-Fernandez J, Luijk R, et al. Genomic and phenotypic insights from an atlas of genetic effects on DNA methylation. Nat Genet. 2021 Sep;53(9):1311–21.

10. Pierce BL, Tong L, Argos M, Demanelis K, Jasmine F, Rakibuz-Zaman M, et al. Co-occurring expression and methylation QTLs allow detection of common causal variants and shared biological mechanisms. Nature Communications [Internet]. 2018 [cited 2022 Jun 6];9. Available from: https://www.ncbi.nlm.nih.gov/pmc/articles/PMC5824840/

11. Kikas T, Rull K, Beaumont RN, Freathy RM, Laan M. The Effect of Genetic Variation on the Placental Transcriptome in Humans. Front Genet. 2019 Jun 11;10:550.

12. Delahaye F, Do C, Kong Y, Ashkar R, Salas M, Tycko B, et al. Genetic variants influence on the placenta regulatory landscape. PLOS Genetics. 2018 Nov 19;14(11):e1007785.

13. Tekola-Ayele F, Zeng X, Chatterjee S, Ouidir M, Lesseur C, Hao K, et al. Placental multi-omics integration identifies candidate functional genes for birthweight. Nat Commun. 2022 May 2;13(1):2384.

14. Bhattacharya A, Li Y, Love MI. MOSTWAS: Multi-Omic Strategies for Transcriptome-Wide Association Studies. PLOS Genetics. 2021 Mar 8;17(3):e1009398.

15. Peng S, Deyssenroth MA, Di Narzo AF, Lambertini L, Marsit CJ, Chen J, et al. Expression quantitative trait loci (eQTLs) in human placentas suggest developmental origins of complex diseases. Hum Mol Genet. 2017 Sep 1;26(17):3432–41.

16. Peng S, Deyssenroth MA, Narzo AFD, Cheng H, Zhang Z, Lambertini L, et al. Genetic regulation of the placental transcriptome underlies birth weight and risk of childhood obesity. PLOS Genetics. 2018 Dec 31;14(12):e1007799.

17. Appleton AA, Murphy MA, Koestler DC, Lesseur C, Paquette AG, Padbury JF, et al. Prenatal Programming of Infant Neurobehaviour in a Healthy Population. Paediatr Perinat Epidemiol. 2016 Jul;30(4):367–75.

18. Oliva M, Muñoz-Aguirre M, Kim-Hellmuth S, Wucher V, Gewirtz ADH, Cotter DJ, et al. The impact of sex on gene expression across human tissues. Science [Internet]. 2020 Sep 11 [cited 2020 Sep 15];369(6509). Available from: https://science.sciencemag.org/content/369/6509/eaba3066

19. Martin E, Smeester L, Bommarito PA, Grace MR, Boggess K, Kuban K, et al. Sexual epigenetic dimorphism in the human placenta: implications for susceptibility during the prenatal period. Epigenomics. 2017 Mar;9(3):267–78.

20. Inkster AM, Yuan V, Konwar C, Matthews AM, Brown CJ, Robinson WP. A cross-cohort analysis of autosomal DNA methylation sex differences in the term placenta. Biology of Sex Differences. 2021 May 27;12(1):38.

21. Bulik-Sullivan BK, Loh PR, Finucane HK, Ripke S, Yang J, Patterson N, et al. LD Score regression distinguishes confounding from polygenicity in genome-wide association studies. Nature Genetics. 2015 Mar 2;47(3):291–5.

22. Finucane HK, Bulik-Sullivan B, Gusev A, Trynka G, Reshef Y, Loh PR, et al. Partitioning heritability by functional annotation using genome-wide association summary statistics. Nature genetics. 2015 Nov;47(11):1228–35.

23. Hormozdiari F, Gazal S, van de Geijn B, Finucane HK, Ju CJT, Loh PR, et al. Leveraging molecular quantitative trait loci to understand the genetic architecture of diseases and complex traits. Nature Genetics. 2018 Jul;50(7):1041–7.

24. Qi T, Wu Y, Zeng J, Zhang F, Xue A, Jiang L, et al. Identifying gene targets for brain-related traits using transcriptomic and methylomic data from blood. Nature Communications. 2018 Jun 11;9(1):2282.

25. Newcombe RG. Two-sided confidence intervals for the single proportion: comparison of seven methods. Statistics in Medicine. 1998;17(8):857–72.

26. Newcombe RG. Interval estimation for the difference between independent proportions: comparison of eleven methods. Statistics in Medicine. 1998;17(8):873–90.

27. Wilson EB. Probable Inference, the Law of Succession, and Statistical Inference. Journal of the American Statistical Association. 1927 Jun 1;22(158):209–12.

28. Iotchkova V, Ritchie GRS, Geihs M, Morganella S, Min JL, Walter K, et al. GARFIELD classifies disease-relevant genomic features through integration of functional annotations with association signals. Nat Genet. 2019 Feb 1;51(2):343–53.

29. Davis CA, Hitz BC, Sloan CA, Chan ET, Davidson JM, Gabdank I, et al. The Encyclopedia of DNA elements (ENCODE): data portal update. Nucleic Acids Res. 2018 04;46(D1):D794–801.

30. Kundaje A, Meuleman W, Ernst J, Bilenky M, Yen A, Heravi-Moussavi A, et al. Integrative analysis of 111 reference human epigenomes. Nature. 2015 Feb;518(7539):317–30.

31. An Integrated Encyclopedia of DNA Elements in the Human Genome. Nature. 2012 Sep 6;489(7414):57–74.

32. Bernstein BE, Stamatoyannopoulos JA, Costello JF, Ren B, Milosavljevic A, Meissner A, et al. The NIH Roadmap Epigenomics Mapping Consortium. Nat Biotechnol. 2010 Oct;28(10):1045–8.

33. Ernst J, Kellis M. Chromatin-state discovery and genome annotation with ChromHMM. Nat Protoc. 2017 Dec;12(12):2478–92.

34. Karmodiya K, Krebs AR, Oulad-Abdelghani M, Kimura H, Tora L. H3K9 and H3K14 acetylation co-occur at many gene regulatory elements, while H3K14ac marks a subset of inactive inducible promoters in mouse embryonic stem cells. BMC Genomics. 2012 Aug 24;13(1):424.

35. Wang Z, Zang C, Rosenfeld JA, Schones DE, Barski A, Cuddapah S, et al. Combinatorial patterns of histone acetylations and methylations in the human genome. Nat Genet. 2008 Jul;40(7):897–903.

36. Elliott G, Hong C, Xing X, Zhou X, Li D, Coarfa C, et al. Intermediate DNA methylation is a conserved signature of genome regulation. Nat Commun. 2015 Feb 18;6(1):6363.

37. Del Gobbo GF, Konwar C, Robinson WP. The significance of the placental genome and methylome in fetal and maternal health. Hum Genet. 2020 Sep 1;139(9):1183–96.

38. Konwar C, Del Gobbo G, Yuan V, Robinson WP. Considerations when processing and interpreting genomics data of the placenta. Placenta. 2019 Sep 1;84:57–62.

39. Bommarito PA, Fry RC. Chapter 2-1 - The Role of DNA Methylation in Gene Regulation. In: McCullough SD, Dolinoy DC, editors. Toxicoepigenetics [Internet]. Academic Press; 2019 [cited 2020 Nov 5]. p. 127–51. Available from: http://www.sciencedirect.com/science/article/pii/B9780128124338000058

40. Edgar R, Tan PPC, Portales-Casamar E, Pavlidis P. Meta-analysis of human methylomes reveals stably methylated sequences surrounding CpG islands associated with high gene expression. Epigenetics & Chromatin. 2014 Oct 23;7(1):28.

41. Gaunt TR, Shihab HA, Hemani G, Min JL, Woodward G, Lyttleton O, et al. Systematic identification of genetic influences on methylation across the human life course. Genome Biology. 2016 Mar 31;17(1):61.

42. Hannon E, Spiers H, Viana J, Pidsley R, Burrage J, Murphy TM, et al. Methylation QTLs in the developing brain and their enrichment in schizophrenia risk loci. Nat Neurosci. 2016 Jan;19(1):48–54.

43. Solmi M, Radua J, Olivola M, Croce E, Soardo L, Salazar de Pablo G, et al. Age at onset of mental disorders worldwide: large-scale meta-analysis of 192 epidemiological studies. Mol Psychiatry. 2021 Jun 2;1–15.

44. Arain M, Haque M, Johal L, Mathur P, Nel W, Rais A, et al. Maturation of the adolescent brain. Neuropsychiatr Dis Treat. 2013;9:449–61.

45. Steinthorsdottir V, McGinnis R, Williams NO, Stefansdottir L, Thorleifsson G, Shooter S, et al. Genetic predisposition to hypertension is associated with preeclampsia in European and Central Asian women. Nat Commun. 2020 Nov 25;11(1):5976.

46. Giambartolomei C, Vukcevic D, Schadt EE, Franke L, Hingorani AD, Wallace C, et al. Bayesian Test for Colocalisation between Pairs of Genetic Association Studies Using Summary Statistics. PLOS Genetics. 2014 May 15;10(5):e1004383.

47. Pardiñas AF, Holmans P, Pocklington AJ, Escott-Price V, Ripke S, Carrera N, et al. Common schizophrenia alleles are enriched in mutation-intolerant genes and in regions under strong background selection. Nature Genetics. 2018 Mar;50(3):381–9.

48. Chowdhury NU, Guntur VP, Newcomb DC, Wechsler ME. Sex and gender in asthma. European Respiratory Review [Internet]. 2021 Dec 31 [cited 2022 Oct 24];30(162). Available from: https://err.ersjournals.com/content/30/162/210067

49. Ferreira MAR, Mathur R, Vonk JM, Szwajda A, Brumpton B, Granell R, et al. Genetic Architectures of Childhood- and Adult-Onset Asthma Are Partly Distinct. Am J Hum Genet. 2019 Apr 4;104(4):665–84.

50. Pividori M, Schoettler N, Nicolae DL, Ober C, Im HK. Shared and Distinct Genetic Risk Factors for Childhood Onset and Adult Onset Asthma: Genome- and Transcriptome-wide Studies. Lancet Respir Med. 2019 Jun;7(6):509–22.

51. Schoettler N, Rodríguez E, Weidinger S, Ober C. Advances in Asthma and Allergic Disease Genetics – Is Bigger Always Better? J Allergy Clin Immunol. 2019 Dec;144(6):1495–506.

52. Kuleshov MV, Jones MR, Rouillard AD, Fernandez NF, Duan Q, Wang Z, et al. Enrichr: a comprehensive gene set enrichment analysis web server 2016 update. Nucleic Acids Res. 2016 Jul 8;44(Web Server issue):W90–7.

53. Xie Z, Bailey A, Kuleshov MV, Clarke DJB, Evangelista JE, Jenkins SL, et al. Gene Set Knowledge Discovery with Enrichr. Current Protocols. 2021;1(3):e90.

54. Chen EY, Tan CM, Kou Y, Duan Q, Wang Z, Meirelles GV, et al. Enrichr: interactive and collaborative HTML5 gene list enrichment analysis tool. BMC Bioinformatics. 2013 Apr 15;14:128.

55. Loisel DA, Tan Z, Tisler CJ, Evans MD, Gangnon RE, Jackson DJ, et al. IFNG genotype and sex interact to influence the risk of childhood asthma. J Allergy Clin Immunol. 2011 Sep;128(3):524–31.

56. Gruen JR, Weissman SM. Human MHC class III and IV genes and disease associations. Frontiers in Bioscience-Landmark. 2001 Aug 1;6(3):960–72.

57. Safran M, Rosen N, Twik M, BarShir R, Stein TI, Dahary D, et al. The GeneCards Suite. In: Abugessaisa I, Kasukawa T, editors. Practical Guide to Life Science Databases [Internet]. Singapore: Springer; 2021 [cited 2022 May 3]. p. 27–56. Available from: https://doi.org/10.1007/978-981-16-5812-9_2

58. Yan Q, Forno E, Herrera-Luis E, Pino-Yanes M, Yang G, Oh S, et al. A genome-wide association study of asthma hospitalizations in adults. Journal of Allergy and Clinical Immunology. 2021 Mar 1;147(3):933–40.

59. Barker DJP, Thornburg KL. The Obstetric Origins of Health for a Lifetime. Clinical Obstetrics & Gynecology. 2013 Sep;56(3):511–9.

60. Bhattacharya A, Freedman AN, Avula V, Harris R, Liu W, Pan C, et al. Placental genomics mediates genetic associations with complex health traits and disease. Nat Commun. 2022 Feb 4;13(1):706.

61. Randall JC, Winkler TW, Kutalik Z, Berndt SI, Jackson AU, Monda KL, et al. Sex-stratified Genome-wide Association Studies Including 270,000 Individuals Show Sexual Dimorphism in Genetic Loci for Anthropometric Traits. PLOS Genetics. 2013 Jun 6;9(6):e1003500.

62. Bernabeu E, Canela-Xandri O, Rawlik K, Talenti A, Prendergast J, Tenesa A. Sex differences in genetic architecture in the UK Biobank. Nat Genet. 2021 Sep;53(9):1283–9.

63. Kautzky-Willer A, Harreiter J, Pacini G. Sex and Gender Differences in Risk, Pathophysiology and Complications of Type 2 Diabetes Mellitus. Endocr Rev. 2016 Jun;37(3):278–316.

64. Alur P. Sex Differences in Nutrition, Growth, and Metabolism in Preterm Infants. Front Pediatr [Internet]. 2019 [cited 2020 Sep 14];7. Available from: https://www.frontiersin.org/articles/10.3389/fped.2019.00022/full

65. Genetic regulation of DNA methylation across tissues reveals thousands of molecular links to complex traits [Internet]. 2021 [cited 2022 Oct 24]. Available from: https://www.researchsquare.com

66. Zhang P, Lewinger JP, Conti D, Morrison JL, Gauderman WJ. Detecting Gene-Environment Interactions for a Quantitative Trait in a Genome-Wide Association Study. Genet Epidemiol. 2016 Jul;40(5):394–403.

67. Martin J, Khramtsova EA, Goleva SB, Blokland GAM, Traglia M, Walters RK, et al. Examining Sex-Differentiated Genetic Effects Across Neuropsychiatric and Behavioral Traits. Biological Psychiatry. 2021 Jun 15;89(12):1127–37.

68. Gay NR, Gloudemans M, Antonio ML, Abell NS, Balliu B, Park Y, et al. Impact of admixture and ancestry on eQTL analysis and GWAS colocalization in GTEx. Genome Biology. 2020 Sep 11;21(1):233.

69. Fenton TR, Kim JH. A systematic review and meta-analysis to revise the Fenton growth chart for preterm infants. BMC Pediatrics. 2013 Apr 20;13(1):59.

70. Lam M, Awasthi S, Watson HJ, Goldstein J, Panagiotaropoulou G, Trubetskoy V, et al. RICOPILI: Rapid Imputation for COnsortias PIpeLIne. Bioinformatics. 2020 Feb 1;36(3):930–3.

71. Chang CC, Chow CC, Tellier LC, Vattikuti S, Purcell SM, Lee JJ. Second-generation PLINK: rising to the challenge of larger and richer datasets. Gigascience [Internet]. 2015 Dec 1 [cited 2020 Nov 3];4(1). Available from: http://academic.oup.com/gigascience/article/4/1/s13742-015-0047-8/2707533

72. Jin Y, Schaffer AA, Feolo M, Holmes JB, Kattman BL. GRAF-pop: A Fast Distance-Based Method To Infer Subject Ancestry from Multiple Genotype Datasets Without Principal Components Analysis. G3 (Bethesda). 2019 May 31;9(8):2447–61.

73. Anderson CA, Pettersson FH, Clarke GM, Cardon LR, Morris AP, Zondervan KT. Data quality control in genetic case-control association studies. Nat Protoc. 2010 Sep;5(9):1564–73.

74. Galinsky KJ, Bhatia G, Loh PR, Georgiev S, Mukherjee S, Patterson NJ, et al. Fast Principal-Component Analysis Reveals Convergent Evolution of ADH1B in Europe and East Asia. Am J Hum Genet. 2016 Mar 3;98(3):456–72.

75. Auton A, Abecasis GR, Altshuler DM, Durbin RM, Abecasis GR, Bentley DR, et al. A global reference for human genetic variation. Nature. 2015 Oct;526(7571):68–74.

76. Das S, Forer L, Schönherr S, Sidore C, Locke AE, Kwong A, et al. Next-generation genotype imputation service and methods. Nat Genet. 2016 Oct;48(10):1284–7.

77. Aryee MJ, Jaffe AE, Corrada-Bravo H, Ladd-Acosta C, Feinberg AP, Hansen KD, et al. Minfi: A flexible and comprehensive Bioconductor package for the analysis of Infinium DNA Methylation microarrays. Bioinformatics. 2014;30(10):1363–9.

78. Chen Y an, Lemire M, Choufani S, Butcher DT, Grafodatskaya D, Zanke BW, et al. Discovery of cross-reactive probes and polymorphic CpGs in the Illumina Infinium HumanMethylation450 microarray. Epigenetics. 2013 Feb 1;8(2):203–9.

79. Price EM, Robinson WP. Adjusting for Batch Effects in DNA Methylation Microarray Data, a Lesson Learned. Frontiers in Genetics [Internet]. 2018 [cited 2020 May 4];9. Available from: http://www.ncbi.nlm.nih.gov/pmc/articles/PMC5864890/

80. Andrey A. Shabalin. Matrix eQTL: ultra fast eQTL analysis via large matrix operations. Bioinformatics. 2012 May 15;28(10):1353–8.

81. Bookman EB, McAllister K, Gillanders E, Wanke K, Balshaw D, Rutter J, et al. Gene-Environment Interplay in Common Complex Diseases: Forging an Integrative Model—Recommendations From an NIH Workshop. Genet Epidemiol. 2011 May;35(4):217–25.

82. Assary E, Vincent JP, Keers R, Pluess M. Gene-environment interaction and psychiatric disorders: Review and future directions. Seminars in Cell & Developmental Biology. 2018 May 1;77:133–43.

83. Do C, Lang CF, Lin J, Darbary H, Krupska I, Gaba A, et al. Mechanisms and Disease Associations of Haplotype-Dependent Allele-Specific DNA Methylation. Am J Hum Genet. 2016 May 5;98(5):934–55.

84. Mostafavi S, Battle A, Zhu X, Urban AE, Levinson D, Montgomery SB, et al. Normalizing RNA-Sequencing Data by Modeling Hidden Covariates with Prior Knowledge. PLoS One [Internet]. 2013 Jul 18 [cited 2020 Mar 3];8(7). Available from: https://www.ncbi.nlm.nih.gov/pmc/articles/PMC3715474/

85. Ng B, White CC, Klein HU, Sieberts SK, McCabe C, Patrick E, et al. An xQTL map integrates the genetic architecture of the human brain’s transcriptome and epigenome. Nature Neuroscience. 2017 Sep 4;20(10):1418–26.

86. Yuan V, Hui D, Yin Y, Peñaherrera M, Beristain A, Robinson W. Cell-specific Characterization of the Placental Methylome [Internet]. In Review; 2020 Oct [cited 2020 Nov 18]. Available from: https://www.researchsquare.com/article/rs-38223/v3

87. Storey JD, Tibshirani R. Statistical significance for genomewide studies. PNAS. 2003 Aug 5;100(16):9440–5.

88. Hannon E, Spiers H, Viana J, Pidsley R, Burrage J, Murphy TM, et al. Methylation quantitative trait loci in the developing brain and their enrichment in schizophrenia-associated genomic regions. Nat Neurosci. 2016 Jan;19(1):48–54.

89. Gazal S, Finucane HK, Furlotte NA, Loh PR, Palamara PF, Liu X, et al. Linkage disequilibrium–dependent architecture of human complex traits shows action of negative selection. Nat Genet. 2017 Oct;49(10):1421–7.

90. Bulik-Sullivan B, Finucane HK, Anttila V, Gusev A, Day FR, Loh PR, et al. An atlas of genetic correlations across human diseases and traits. Nat Genet. 2015 Nov;47(11):1236–41.

91. Hormozdiari F, van de Bunt M, Segrè AV, Li X, Joo JWJ, Bilow M, et al. Colocalization of GWAS and eQTL Signals Detects Target Genes. Am J Hum Genet. 2016 Dec 1;99(6):1245–60.

92. Lumley T. rmeta: Meta-Analysis [Internet]. 2018. Available from: https://CRAN.R-project.org/package=rmeta

93. Kent WJ, Sugnet CW, Furey TS, Roskin KM, Pringle TH, Zahler AM, et al. The Human Genome Browser at UCSC. Genome Res. 2002 Jan 6;12(6):996–1006.

