## Supplementary Figures and Small Supplementary Tables for "Sex-dependent placental mQTL provide insight into the prenatal origins of childhood-onset traits and conditions"

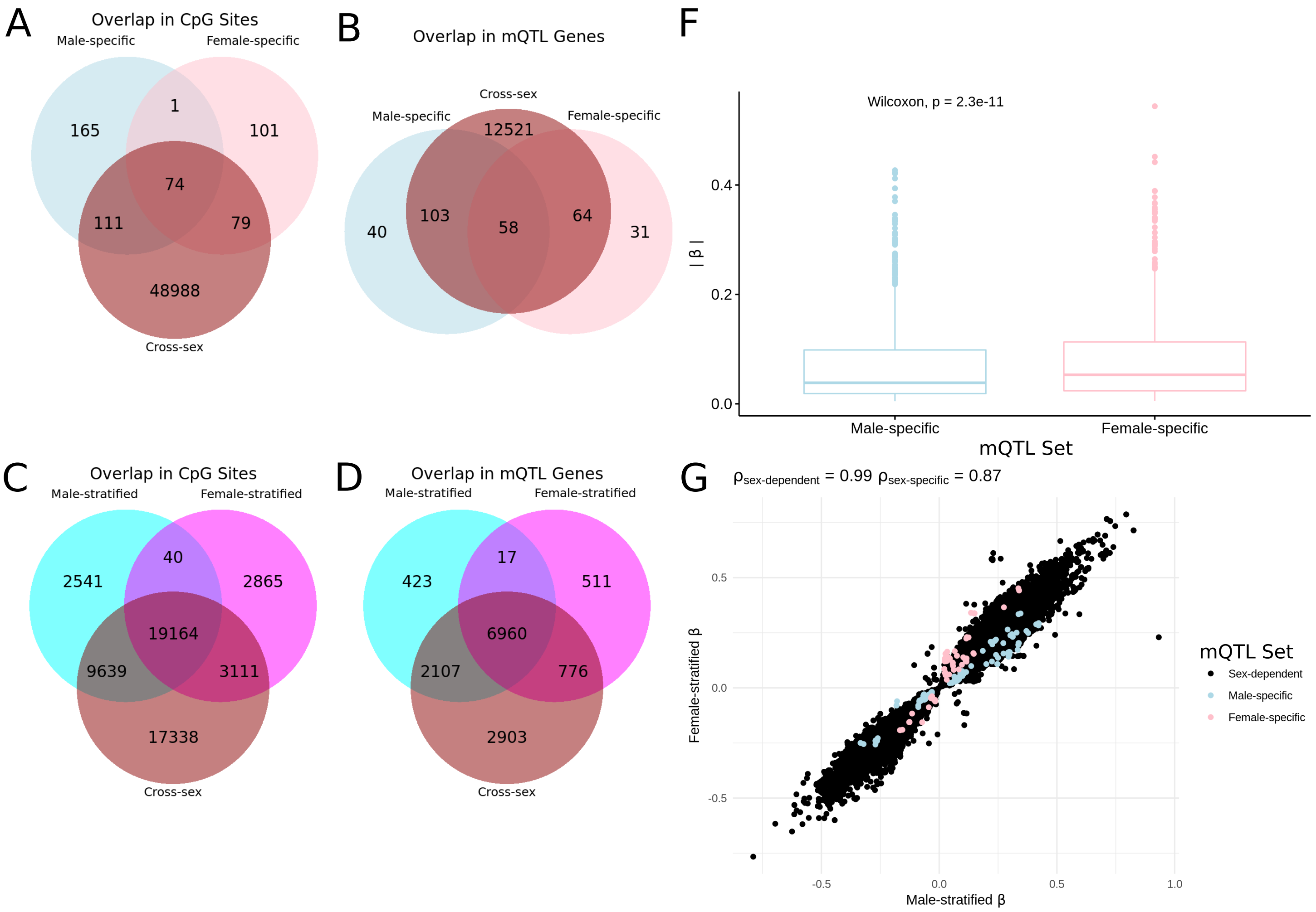


**Supplementary Figure S1. Summary of CpG counts, gene counts, and effect sizes for all mQTL** CpG sites with at least one mQTL. We present CpG sites with at least one cross-sex, male- or female-specific mQTL (A), and counts for genes on the Illumina450kArray annotation targeted by these CpG sites (B). Next, we present the same measures for CpG sites with at least one cross-sex, male- or female-stratified mQTL (C), and the genes targeted by these CpG sites (D). CpG sites shared across mQTL sets do not necessarily reflect the same mQTL, as different SNPs from each category can be associated with DNAm at the same CpG site. We then compare effect sizes for male- and female-specific placental mQTL. (F) Differences in the absolute male- and female-stratified mQTL effect size for male- and female-specific mQTL and show the two-sided Wilcoxon rank-sum test p value(mean absolute beta 0.067 in males vs. 0.079 in Females, $p < 2.3e\text{-}11$). (G)Male- vs. female-stratified mQTL effect sizes for all sex-dependent mQTL, male- and female-specific mQTL are colored. Although differences between male- vs. female-stratified effect sizes are small for these male- and female-specific mQTL, note that they have significant interaction effects (i.e., they are sex-dependent). Correlation between male- and female-stratified effects is marked "sex-dependent" (within sex-dependent mQTL) or as "sex-specific" (within sex-dependent mQTL with either a male- or female-specific effect).

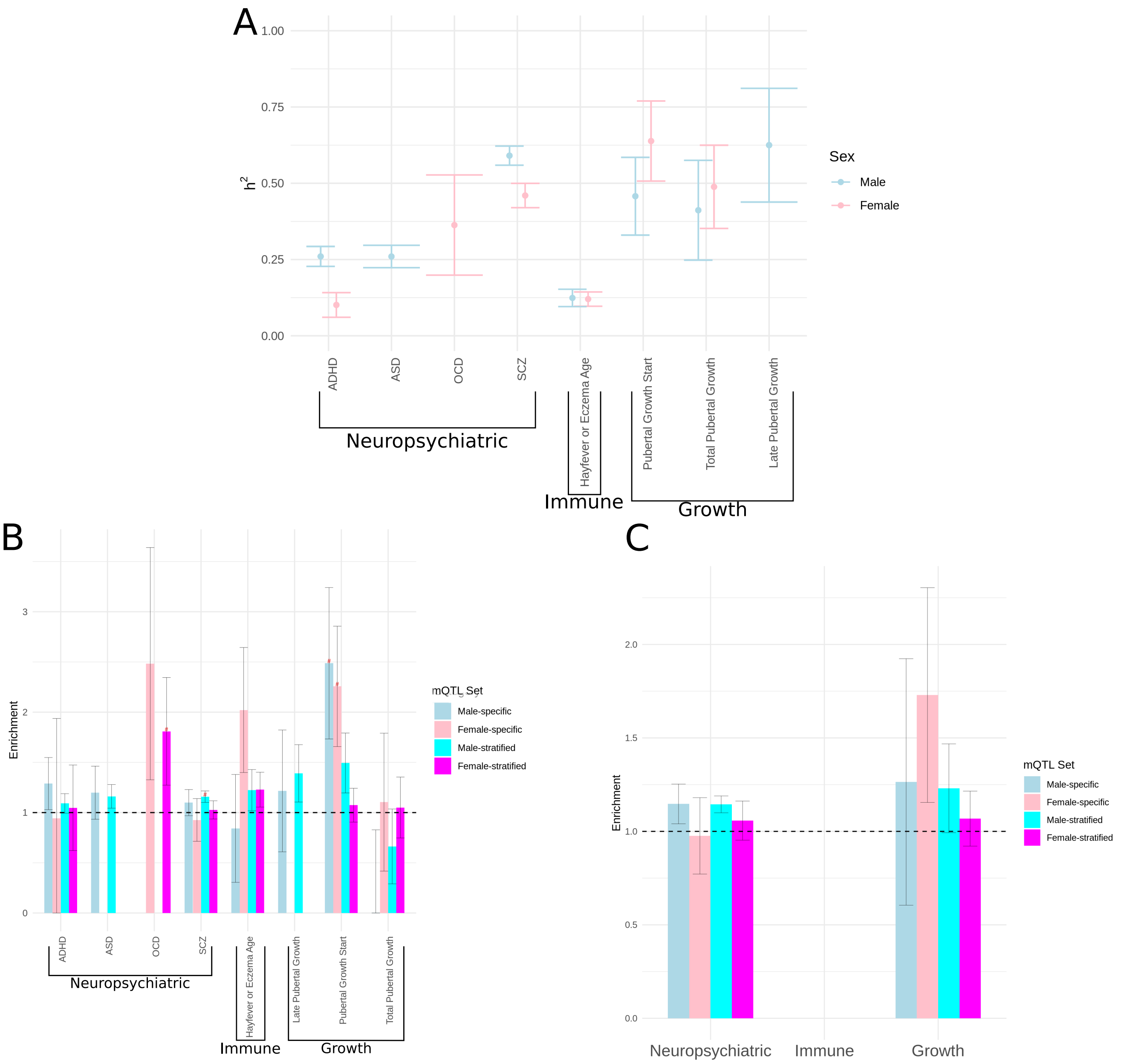


**Supplementary Figure S2. Stratified linkage disequilibrium score regression in male- and female-stratified GWAS summary statistics.** (A) $h^2_{SNP}$ estimates for each male- and female-stratified GWAS that pass quality control along with their standard error. (B) Male- and female-specific and male- and female-stratified mQTL S-LDSC enrichment in male- and female-stratified GWAS respectively. (C) Random-effect meta-analysis of S-LDSC enrichment within three trait categories: neuropsychiatric-, immune-, and growth-related. Estimates with a within-trait FDR $<$ 0.05, are marked with a "#", whereas estimates with an FDR $<$ 0.05 accounting for all tests are marked with an asterisk. Estimating significance for meta-analyzed S-LDSC estimates is not defined and is thus not shown here.

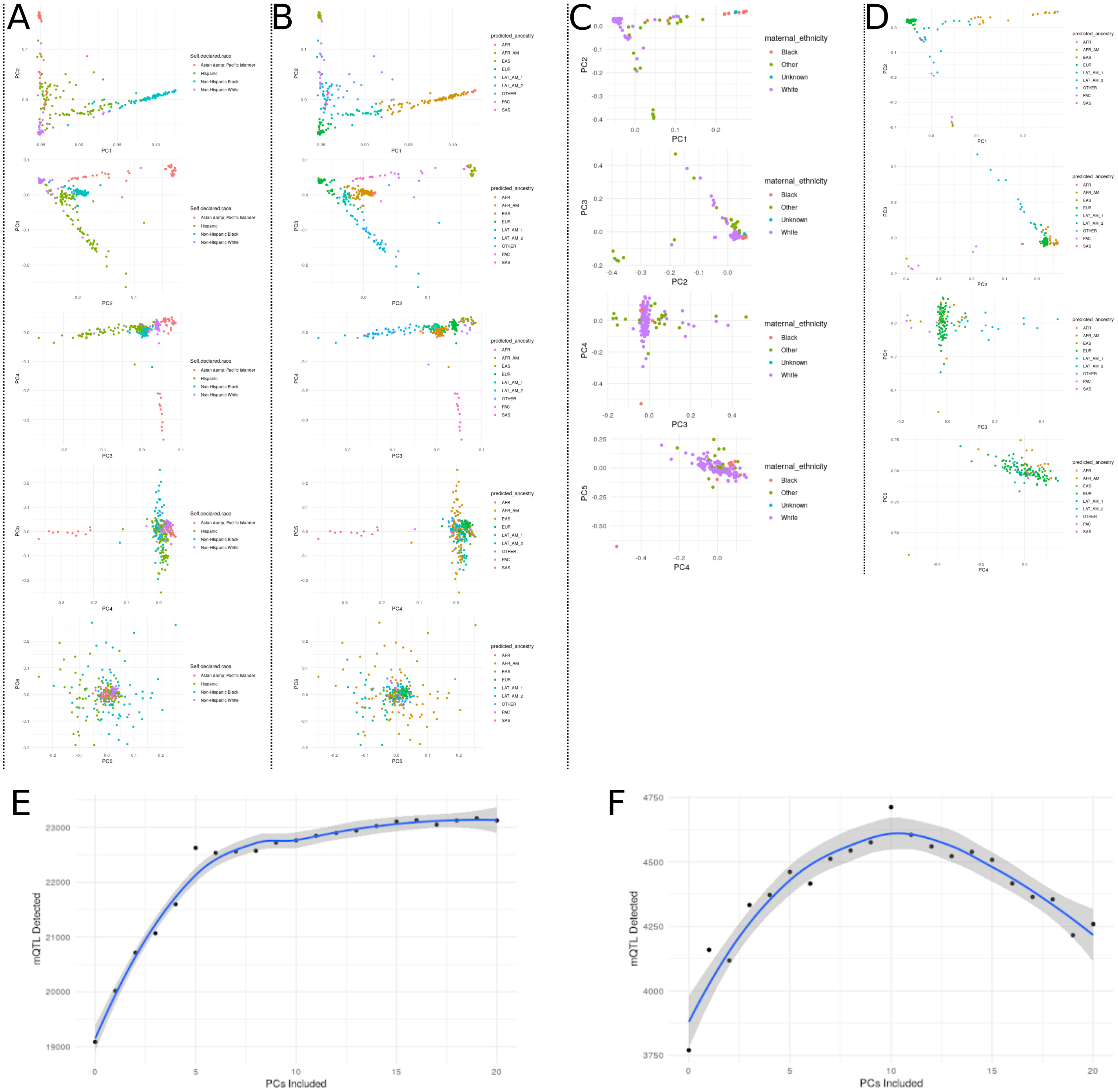


**Supplementary Figure S3. Developing genotyping and DNAm derived covariates for mQTL analyses in NICHD and RICHS.** Samples in NICHD plotted by first six ancestry PCs, colored by self-reported maternal ethnicity (A) and GRAF-pop estimated ancestry (B). Samples in RICHS colored by their first five ancestry PCs, again colored by self-reported maternal ethnicity (C) and GRAF-pop estimated ancestry (D). Genome-wide principal components of DNAm regressed on mQTL in Chromosome 21 in order of variance explained vs. the number of mQTL detected at a Bonferroni corrected $p < 0.05$ for all SNPs within 75kb of each CpG site in NICHD samples (E) and RICHS samples (F).

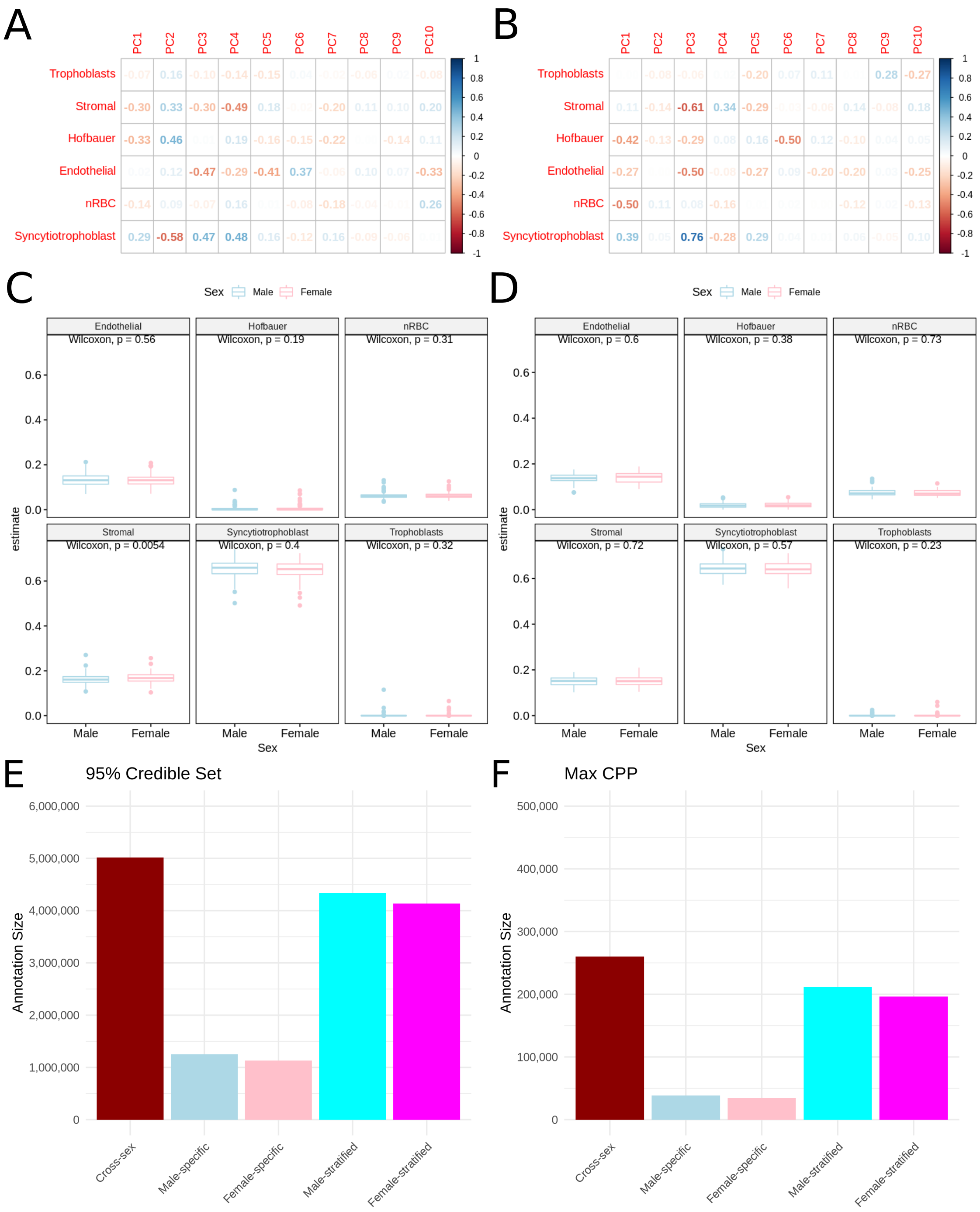


**Supplementary Figure S4. Assessing limitations of mQTL models**. The correlation between estimated cell type proportions and the first 10 DNAm PCs in NICHD (A) and RICHS (B). Estimated cell type proportions of samples passing quality control in male vs. female participants in NICHD (C) and RICHS (D). A significant difference (nominal two-sided Wilcox test $p < 0.05$) in estimated stromal cell proportion was found in NICHD, and no significant differences between male vs. female cell type proportions were detected in RICHS. The sum over the 95% credible set (E) and maximum causal posterior probability annotations (F) for five mQTL sets.

**Supplementary Table S1. Population level summaries of the NICHD and RICHS studies by sex.** Percentages and standard deviations (SD) are taken with respect to each sex within each sample, with the exception of the Number of Subjects, where the percentage reported is with respect to each sample. Measurements that were not reported for a given study are labelled with NA.

| | NICHD Samples | | RICHS Samples | |
|---|---|---|---|---|
| | Males | Females | Males | Females |
| Number of Subjects N (%) | 149 (54.2) | 126 (45.8) | 73 (53.6) | 63 (46.3) |
| Mean Gestational Age in Weeks (SD) | 39.4 (1.2) | 39.6 (1.0) | 39.1 (0.95) | 39.0 ( 0.96) |
| Small for Gestational Age Fenton Group N (%) | NA | NA | 8 (11.0) | 13 (20.6) |
| Average for Gestational Age Fenton Group N (%) | NA | NA | 40 (54.8) | 35 (55.6) |
| Large for Gestational Age Fenton Group N (%) | NA | NA | 25 (39.7) | 15 (20.5) |
| Pre-eclampsia Cases N (%) | 5 (3.4) | 4 (3.2) | NA | NA |
| Mean Birth Weight in Grams (SD) | 3183.7 (480.5) | 3126.3 (404.0) | NA | NA |
| Reported Maternal Ethnicity | | | | |
| Asian & Pacific Islander N (%) | 31 (24.6) | 10 (7.9) | NA | NA |
| Hispanic N (%) | 47 (31.6) | 44 (29.5) | NA | NA |
| Non-Hispanic Black N (%) | 33 (26.2) | 40 (31.7) | 4 (5.5) | 6 (9.5) |
| White N (%) | 38 (25.5) | 32 (21.5) | 60 (82.2) | 43 (58.9) |
| Other | NA | NA | 9 (12.3) | 12 (19.0) |
| Unknown | NA | NA | 0 (0.0) | 2 (3.2) |

**Supplementary Table S2. Differences in proportion of CpG sites with at least one mQTL from six categories.** The proportion of CpG sites with at least one mQTL in each chromosome relative to all chromosomes for each mQTL set. We show the differences in these proportions across mQTL categories using the proportion test. Differences were declared significant at Holm-Bonferroni corrected $p < 0.05$ within each set of proportions.

| Chr | Cross-sex probes | Sex-dependent probes | Cross-sex vs sex-dependent proportion difference | Cross-sex vs sex-dependent p | Cross-sex vs sex-dependent adjusted p | Male-specific CpGs | Female-specific CpGs | Male- vs female-specific proportion difference | Male- vs female-specific p | Male- vs female-specific adjusted p | Male-stratified CpGs | Female-stratified CpGs | Male- vs female-stratified proportion difference | Male- vs female-stratified p | Male- vs female-stratified adjusted p |
|---|---|---|---|---|---|---|---|---|---|---|---|---|---|---|---|
| 1 | 4671 | 249 | 5.20E-03 | 4.08E-01 | 5.21E-01 | 54 | 31 | 3.23E-02 | 3.12E-01 | 8.97E-01 | 2957 | 2345 | 1.09E-03 | 6.69E-01 | 1 |
| 2 | 3542 | 167 | 4.82E-03 | 3.84E-01 | 5.20E-01 | 21 | 11 | 1.67E-02 | 4.70E-01 | 9.82E-01 | 2265 | 1774 | 1.72E-03 | 4.40E-01 | 9.44E-01 |
| 3 | 2364 | 136 | 6.64E-03 | 1.44E-01 | 2.37E-01 | 16 | 20 | 3.28E-02 | 1.30E-01 | 5.39E-01 | 1523 | 1217 | 1.96E-04 | 9.30E-01 | 1 |
| 4 | 2060 | 99 | 2.05E-03 | 6.54E-01 | 7.52E-01 | 12 | 10 | 5.03E-03 | 9.15E-01 | 1 | 1339 | 1075 | 2.76E-05 | 1 | 1 |
| 5 | 2333 | 125 | 2.85E-03 | 5.46E-01 | 6.60E-01 | 9 | 17 | 4.10E-02 | 2.40E-02 | 1.48E-01 | 1474 | 1192 | 3.73E-04 | 8.51E-01 | 1 |
| 7 | 3239 | 163 | 2.76E-04 | 9.90E-01 | 9.90E-01 | 19 | 9 | 1.88E-02 | 3.71E-01 | 9.48E-01 | 3252 | 2600 | 3.63E-04 | 8.99E-01 | 1 |
| 9 | 995 | 37 | 5.34E-03 | 7.43E-02 | 1.32E-01 | 19 | 22 | 3.21E-02 | 1.64E-01 | 5.39E-01 | 2087 | 1650 | 9.71E-04 | 6.56E-01 | 1 |
| 10 | 2622 | 111 | 8.64E-03 | 6.66E-02 | 1.32E-01 | 7 | 11 | 2.32E-02 | 1.56E-01 | 5.39E-01 | 1408 | 1164 | 1.36E-03 | 4.51E-01 | 9.44E-01 |
| 13 | 1226 | 51 | 4.40E-03 | 1.89E-01 | 2.89E-01 | 2 | 3 | 6.07E-03 | 7.19E-01 | 1 | 634 | 485 | 9.40E-04 | 4.43E-01 | 9.44E-01 |
| 14 | 1355 | 65 | 1.40E-03 | 7.24E-01 | 7.93E-01 | 20 | 11 | 1.38E-02 | 5.64E-01 | 1 | 1733 | 1396 | 2.22E-04 | 9.23E-01 | 1 |
| 15 | 1385 | 79 | 3.62E-03 | 3.17E-01 | 4.56E-01 | 11 | 19 | 4.32E-02 | 2.58E-02 | 1.48E-01 | 1852 | 1650 | 6.52E-03 | 1.48E-03 | 1.70E-02 |
| 16 | 2412 | 120 | 7.60E-04 | 9.01E-01 | 9.42E-01 | 17 | 11 | 5.30E-03 | 9.12E-01 | 1 | 1401 | 1177 | 2.10E-03 | 2.41E-01 | 9.44E-01 |
| 22 | 237 | 19 | 2.82E-03 | 7.01E-02 | 1.32E-01 | 2 | 2 | 2.15E-03 | 1 | 1 | 739 | 594 | 4.31E-05 | 9.95E-01 | 1 |
| 6 | 4942 | 112 | 5.53E-02 | 1.58E-19 | 1.81E-18 | 17 | 3 | 3.67E-02 | 2.36E-02 | 1.48E-01 | 903 | 718 | 2.58E-04 | 8.75E-01 | 1 |
| 8 | 2259 | 74 | 1.61E-02 | 1.87E-04 | 8.62E-04 | 8 | 8 | 8.58E-03 | 6.94E-01 | 1 | 892 | 777 | 2.44E-03 | 9.37E-02 | 7.18E-01 |
| 11 | 2934 | 182 | 1.36E-02 | 6.35E-03 | 1.82E-02 | 18 | 10 | 1.21E-02 | 6.15E-01 | 1 | 1524 | 1184 | 1.54E-03 | 4.06E-01 | 9.44E-01 |
| 12 | 2312 | 145 | 1.13E-02 | 1.10E-02 | 2.54E-02 | 24 | 16 | 5.63E-03 | 9.12E-01 | 1 | 1884 | 1466 | 1.81E-03 | 3.74E-01 | 9.44E-01 |
| 17 | 3020 | 185 | 1.30E-02 | 9.75E-03 | 2.49E-02 | 2 | 4 | 9.99E-03 | 4.18E-01 | 9.61E-01 | 288 | 223 | 3.20E-04 | 7.22E-01 | 1 |
| 18 | 483 | 42 | 7.07E-03 | 8.68E-04 | 2.85E-03 | 14 | 12 | 7.17E-03 | 8.20E-01 | 1 | 1349 | 1097 | 5.83E-04 | 7.51E-01 | 1 |
| 19 | 982 | 160 | 4.43E-02 | 2.03E-48 | 4.68E-47 | 16 | 10 | 6.37E-03 | 8.58E-01 | 1 | 615 | 528 | 1.37E-03 | 2.61E-01 | 9.44E-01 |
| 20 | 1032 | 78 | 1.04E-02 | 6.31E-04 | 2.42E-03 | 9 | 5 | 6.03E-03 | 8.30E-01 | 1 | 363 | 286 | 2.08E-04 | 8.48E-01 | 1 |
| 21 | 2265 | 28 | 3.47E-02 | 3.18E-16 | 2.44E-15 | 3 | 2 | 7.04E-04 | 1 | 1 | 632 | 554 | 1.86E-03 | 1.32E-01 | 7.56E-01 |
| X | 582 | 62 | 1.31E-02 | 1.55E-08 | 8.93E-08 | 31 | 8 | 5.69E-02 | 7.99E-03 | 1.48E-01 | 270 | 28 | 7.49E-03 | 4.34E-34 | 9.99E-33 |

**Supplementary Table S3. Difference in mean signed and absolute effect size for male- and female-specific mQTL.** p values are from a two-sided Wilcoxon rank-sum test between male and female effects. Holm-Bonferroni adjusted p values were computed with respect to signed and absolute effect sizes respectfully.

| Chr | Mean beta in male-specific mQTL | Mean beta in female-specific mQTL | p beta | Holm-Bonferroni adjusted p beta | Mean absolute beta in male-specific mQTL | Mean absolute beta in female-specific mQTL | p absolute beta | Holm-Bonferroni Adjusted p absolute beta |
|---|---|---|---|---|---|---|---|---|
| 1 | 2.35E-02 | 5.99E-03 | 6.48E-13 | 1.40E-11 | 5.58E-02 | 6.19E-02 | 2.58E-05 | 4.40E-04 |
| 2 | -6.88E-03 | 3.33E-02 | 1.12E-01 | 6.90E-01 | 4.22E-02 | 5.70E-02 | 3.52E-02 | 3.90E-01 |
| 3 | 1.69E-02 | 6.38E-02 | 1.18E-01 | 6.90E-01 | 3.56E-02 | 9.68E-02 | 2.01E-08 | 3.80E-07 |
| 4 | 1.42E-02 | 6.00E-02 | 6.62E-06 | 1.20E-04 | 5.69E-02 | 7.13E-02 | 4.92E-01 | 9.80E-01 |
| 5 | 6.14E-02 | -1.40E-02 | 4.45E-05 | 7.10E-04 | 6.21E-02 | 6.66E-02 | 2.37E-01 | 7.10E-01 |
| 6 | -7.57E-03 | -4.04E-02 | 6.06E-16 | 1.30E-14 | 3.16E-02 | 6.62E-02 | 3.00E-19 | 6.30E-18 |
| 7 | 1.28E-02 | 1.38E-01 | 4.21E-41 | 9.70E-40 | 3.33E-02 | 1.45E-01 | 1.26E-45 | 2.90E-44 |
| 8 | 2.13E-02 | 3.07E-02 | 3.17E-03 | 3.80E-02 | 5.90E-02 | 3.62E-02 | 3.16E-07 | 5.70E-06 |
| 9 | 3.68E-02 | -4.18E-02 | 3.72E-01 | 1.00E+00 | 6.83E-02 | 4.18E-02 | 4.97E-02 | 4.70E-01 |
| 10 | 1.33E-02 | 1.12E-02 | 1.33E-06 | 2.50E-05 | 8.57E-02 | 4.77E-02 | 3.58E-29 | 7.90E-28 |
| 11 | 2.95E-02 | -2.72E-02 | 5.39E-04 | 7.50E-03 | 3.71E-02 | 5.56E-02 | 8.36E-02 | 5.00E-01 |
| 12 | -2.67E-02 | -4.76E-02 | 1.82E-01 | 7.30E-01 | 5.99E-02 | 5.66E-02 | 1.27E-02 | 1.50E-01 |
| 13 | 7.53E-03 | 2.01E-02 | 4.35E-02 | 3.90E-01 | 1.11E-02 | 4.49E-02 | 7.08E-04 | 9.90E-03 |
| 14 | 7.14E-03 | 1.20E-01 | 4.14E-01 | 1.00E+00 | 3.70E-02 | 1.54E-01 | 4.72E-02 | 4.70E-01 |
| 15 | 9.94E-02 | 1.39E-01 | 3.45E-03 | 3.80E-02 | 1.03E-01 | 1.48E-01 | 9.99E-04 | 1.30E-02 |
| 16 | 2.92E-02 | 1.06E-01 | 2.96E-11 | 5.90E-10 | 3.70E-02 | 1.10E-01 | 5.08E-12 | 1.00E-10 |
| 17 | 4.28E-02 | 9.38E-03 | 1.27E-04 | 1.90E-03 | 7.24E-02 | 4.60E-02 | 5.34E-02 | 4.70E-01 |
| 18 | -1.86E-02 | -1.26E-02 | 5.73E-01 | 1.00E+00 | 3.15E-02 | 8.64E-02 | 1.88E-04 | 3.00E-03 |
| 19 | -2.69E-03 | 8.55E-02 | 4.71E-02 | 3.90E-01 | 5.76E-02 | 1.12E-01 | 5.38E-01 | 9.80E-01 |
| 20 | 8.30E-03 | -1.66E-02 | 6.28E-03 | 6.30E-02 | 5.98E-02 | 3.96E-02 | 5.96E-02 | 4.70E-01 |
| 21 | 2.98E-03 | 6.02E-02 | 5.52E-04 | 7.50E-03 | 4.26E-02 | 8.58E-02 | 2.63E-04 | 3.90E-03 |
| 22 | 7.29E-02 | 3.58E-02 | 9.88E-02 | 6.90E-01 | 7.29E-02 | 4.74E-02 | 9.88E-02 | 5.00E-01 |
| X | 6.18E-02 | 1.88E-01 | 1.48E-05 | 2.50E-04 | 1.57E-01 | 1.88E-01 | 1.02E-01 | 5.00E-01 |

**Supplementary Table S4. Random-effect meta-analysis of enrichment values and tau coefficients from stratified LD score regression.** A random-effect meta-analysis of enrichment and $\tau^*$ values from S-LDSC within each mQTL set and across GWAS from one of 3 categories of related traits and conditions: neuropsychiatric-, immune-, and growth-related. GWAS Categories for which more than one sex-stratified GWAS summary statistics were available and passing quality control are labeled _male or _female respectively; only male- or female-stratified GWAS summary statistics were considered in the meta-analysis. GWAS categories with one or fewer GWAS have their effects listed as NA.

| GWAS Category | mQTL Set | Enrichment | Enrichment_std_error | tau_star | tau_se | tau_p |
|---|---|---|---|---|---|---|
| Neuropsychiatric | Cross-sex | 1.02 | 0.0316 | -0.000178 | 0.00105 | 0.865 |
| Neuropsychiatric | Male-specific | 0.973 | 0.115 | -0.000125 | 0.000655 | 0.849 |
| Neuropsychiatric | Female-specific | 0.938 | 0.0868 | -0.000424 | 0.000496 | 0.393 |
| Neuropsychiatric | Male-stratified | 1.01 | 0.0452 | 6.92e-05 | 0.000903 | 0.939 |
| Neuropsychiatric | Female-stratified | 1.01 | 0.0403 | -0.000262 | 0.000887 | 0.768 |
| Neuropsychiatric_male | Male-specific | 1.15 | 0.106 | 0.000383 | 0.000591 | 0.517 |
| Neuropsychiatric_male | Male-stratified | 1.14 | 0.0453 | 0.00232 | 0.00101 | 0.0219 |
| Neuropsychiatric_female | Female-specific | 0.976 | 0.204 | 0.000695 | 0.00229 | 0.762 |
| Neuropsychiatric_female | Female-stratified | 1.06 | 0.105 | 0.00438 | 0.00564 | 0.437 |
| Immune | Cross-sex | 1.29 | 0.0618 | -0.00213 | 0.00243 | 0.38 |
| Immune | Male-specific | 1.49 | 0.221 | -0.000933 | 0.00149 | 0.53 |
| Immune | Female-specific | 1.75 | 0.249 | 0.000593 | 0.00144 | 0.681 |
| Immune | Male-stratified | 1.29 | 0.0855 | -0.00309 | 0.00257 | 0.229 |
| Immune | Female-stratified | 1.28 | 0.0971 | -0.00356 | 0.00259 | 0.169 |
| Immune_male | Male-specific | NA | NA | NA | NA | NA |
| Immune_male | Male-stratified | NA | NA | NA | NA | NA |
| Immune_female | Female-specific | NA | NA | NA | NA | NA |
| Immune_female | Female-stratified | NA | NA | NA | NA | NA |
| Growth | Cross-sex | 1.2 | 0.0831 | 0.00151 | 0.00146 | 0.301 |
| Growth | Male-specific | 1.66 | 0.167 | 0.00158 | 0.000966 | 0.102 |
| Growth | Female-specific | 1.78 | 0.137 | 0.00169 | 0.000743 | 0.0226 |
| Growth | Male-stratified | 1.24 | 0.0775 | 0.00114 | 0.0011 | 0.301 |
| Growth | Female-stratified | 1.24 | 0.0901 | 0.00113 | 0.00115 | 0.325 |
| Growth_male | Male-specific | 1.26 | 0.659 | -0.000735 | 0.00381 | 0.847 |
| Growth_male | Male-stratified | 1.23 | 0.238 | -0.000586 | 0.0066 | 0.929 |
| Growth_female | Female-specific | 1.73 | 0.574 | 0.0024 | 0.00292 | 0.41 |
| Growth_female | Female-stratified | 1.07 | 0.147 | -0.00388 | 0.00332 | 0.242 |

**Supplementary Table S5. Counts of self-reported maternal ethnicity vs. GRAF-pop estimated ancestry for NICHD and RICHS placental samples.** Ancestry was estimated on raw genotypes using GRAF-pop (Star Methods), and we enumerate this vs. each sample's maternal self-reported ancestry in the NICHD and RICHS studies.

| | | Self-Reported Maternal Ethnicity NICHD | | | | Self-Reported Maternal Ethnicity RICHS | | | |
|---|---|---|---|---|---|---|---|---|---|
| | | Asian & Pacific Islander | Hispanic | Non-Hispanic Black | Non-Hispanic White | Black | Other | Unknown | White |
| GRAF-pop Estimated Ancestry | AFR | 0 | 0 | 7 | 0 | 2 | 0 | 0 | 0 |
| | AFR_AM | 0 | 14 | 64 | 4 | 6 | 7 | 2 | 3 |
| | EAS | 27 | 0 | 0 | 0 | 0 | 2 | 0 | 0 |
| | EUR | 0 | 5 | 0 | 67 | 0 | 0 | 0 | 99 |
| | LAT_AM_1 | 0 | 28 | 1 | 1 | 2 | 4 | 0 | 2 |
| | LAT_AM_2 | 0 | 43 | 0 | 3 | 0 | 4 | 0 | 7 |
| | OTHER | 1 | 11 | 1 | 0 | 0 | 0 | 0 | 2 |
| | PAC | 11 | 1 | 0 | 2 | 0 | 4 | 1 | 1 |
| | SAS | 11 | 0 | 0 | 0 | 0 | 0 | 0 | 1 |
